# The relationship between BMI and COVID-19: exploring misclassification and selection bias in a two-sample Mendelian randomisation study

**DOI:** 10.1101/2022.03.03.22271836

**Authors:** Gemma L Clayton, Ana Gonçalves Soares, Neil Goulding, Maria Carolina Borges, Michael V Holmes, George Davey Smith, Kate Tilling, Deborah A Lawlor, Alice R Carter

## Abstract

**Objective:** To use the example of the effect of body mass index (BMI) on COVID-19 susceptibility and severity to illustrate methods to explore potential selection and misclassification bias in Mendelian randomisation (MR) of COVID-19 determinants.

**Design:** Two-sample MR analysis.

**Setting:** Summary statistics from the Genetic Investigation of ANthropometric Traits (GIANT) and COVID-19 Host Genetics Initiative (HGI) consortia.

**Participants:** 681,275 participants in GIANT and more than 2.5 million people from the COVID-19 HGI consortia.

**Exposure:** Genetically instrumented BMI.

**Main outcome measures:** Seven case/control definitions for SARS-CoV-2 infection and COVID-19 severity: very severe respiratory confirmed COVID-19 vs not hospitalised COVID-19 (A1) and vs population (those who were never tested, tested negative or had unknown testing status (A2)); hospitalised COVID-19 vs not hospitalised COVID-19 (B1) and vs population (B2); COVID-19 vs lab/self-reported negative (C1) and vs population (C2); and predicted COVID-19 from self-reported symptoms vs predicted or self-reported non-COVID-19 (D1).

**Results:** With the exception of A1 comparison, genetically higher BMI was associated with higher odds of COVID-19 in all comparison groups, with odds ratios (OR) ranging from 1.11 (95%CI: 0.94, 1.32) for D1 to 1.57 (95%CI: 1.57 (1.39, 1.78) for A2. As a method to assess selection bias, we found no strong evidence of an effect of COVID-19 on BMI in a ‘no-relevance’ analysis, in which COVID-19 was considered the exposure, although measured after BMI. We found evidence of genetic correlation between COVID-19 outcomes and potential predictors of selection determined *a priori* (smoking, education, and income), which could either indicate selection bias or a causal pathway to infection. Results from multivariable MR adjusting for these predictors of selection yielded similar results to the main analysis, suggesting the latter.

**Conclusions:** We have proposed a set of analyses for exploring potential selection and misclassification bias in MR studies of risk factors for SARS-CoV-2 infection and COVID-19 and demonstrated this with an illustrative example. Although selection by socioeconomic position and arelated traits is present, MR results are not substantially affected by selection/misclassification bias in our example. We recommend the methods we demonstrate, and provide detailed analytic code for their use, are used in MR studies assessing risk factors for COVID-19, and other MR studies where such biases are likely in the available data.

**Summary:** *What is already known on this topic:*

- Mendelian randomisation (MR) studies have been conducted to investigate the potential causal relationship between body mass index (BMI) and COVID-19 susceptibility and severity.
- There are several sources of selection (e.g. when only subgroups with specific characteristics are tested or respond to study questionnaires) and misclassification (e.g. those not tested are assumed not to have COVID-19) that could bias MR studies of risk factors for COVID-19.
- Previous MR studies have not explored how selection and misclassification bias in the underlying genome-wide association studies could bias MR results.

*What this study adds:*

- Using the most recent release of the COVID-19 Host Genetics Initiative data (with data up to June 2021), we demonstrate a potential causal effect of BMI on susceptibility to detected SARS-CoV-2 infection and on severe COVID-19 disease, and that these results are unlikely to be substantially biased due to selection and misclassification.
- This conclusion is based on no evidence of an effect of COVID-19 on BMI (a ‘no-relevance control’ study, as BMI was measured before the COVID-19 pandemic) and finding genetic correlation between predictors of selection (e.g. socioeconomic position) and COVID-19 for which multivariable MR supported a role in causing susceptibility to infection.
- We recommend studies use the set of analyses demonstrated here in future MR studies of COVID-19 risk factors, or other examples where selection bias is likely.

## Introduction

Mendelian randomisation (MR) has been used to explore risk factors for disease occurrence and prognosis, including for severe acute respiratory syndrome coronavirus 2 (SARS-CoV-2), the virus that causes coronavirus disease 2019 (COVID-19, hereafter referred to as COVID-19 susceptibility/severity) (1, 2). MR can be implemented as an instrumental variable (IV) analysis, which uses genetic variants related to potential exposures, or modifiable risk factors, to explore their causal effects (3, 4). Because genetic variants are randomly allocated at conception, they are not modified throughout the life course, and therefore MR is less likely to be biased by confounding or prevalent disease (i.e., reverse causality) than conventional multivariable regression (3-5). However, like other study designs, MR can be biased, invalidating inferences made (4, 6). Whilst both horizontal pleiotropy (where genetic variants influence the outcome through pathways not including the risk factor of interest) and confounding by population stratification are widely discussed (7-9), selection and misclassification bias are less commonly discussed, particularly in applied papers (6, 10, 11).

### Selection bias

Selection bias is a systematic error introduced when a study sample systematically differs from the target population, e.g., when there is dropout, subgroup analyses or missing data (12). In the context of COVID-19, selection bias can arise from differential response to COVID-19 study questionnaires, differential risk of being exposed to SARS-Cov-2 infection, and differences in who receives a SARS-CoV-2 test, or who is admitted to hospital (13, 14). For example, if a person never had a SARS-CoV-2 test, then their COVID-19 status is unknown, and they would not be selected into any analysis that depends on having a test (See **Table 1** for more details). Selection bias can occur in genome wide association studies (GWAS) when participants selected into the GWAS are not a random sample of the target population (15, 16).

**Table 1.**
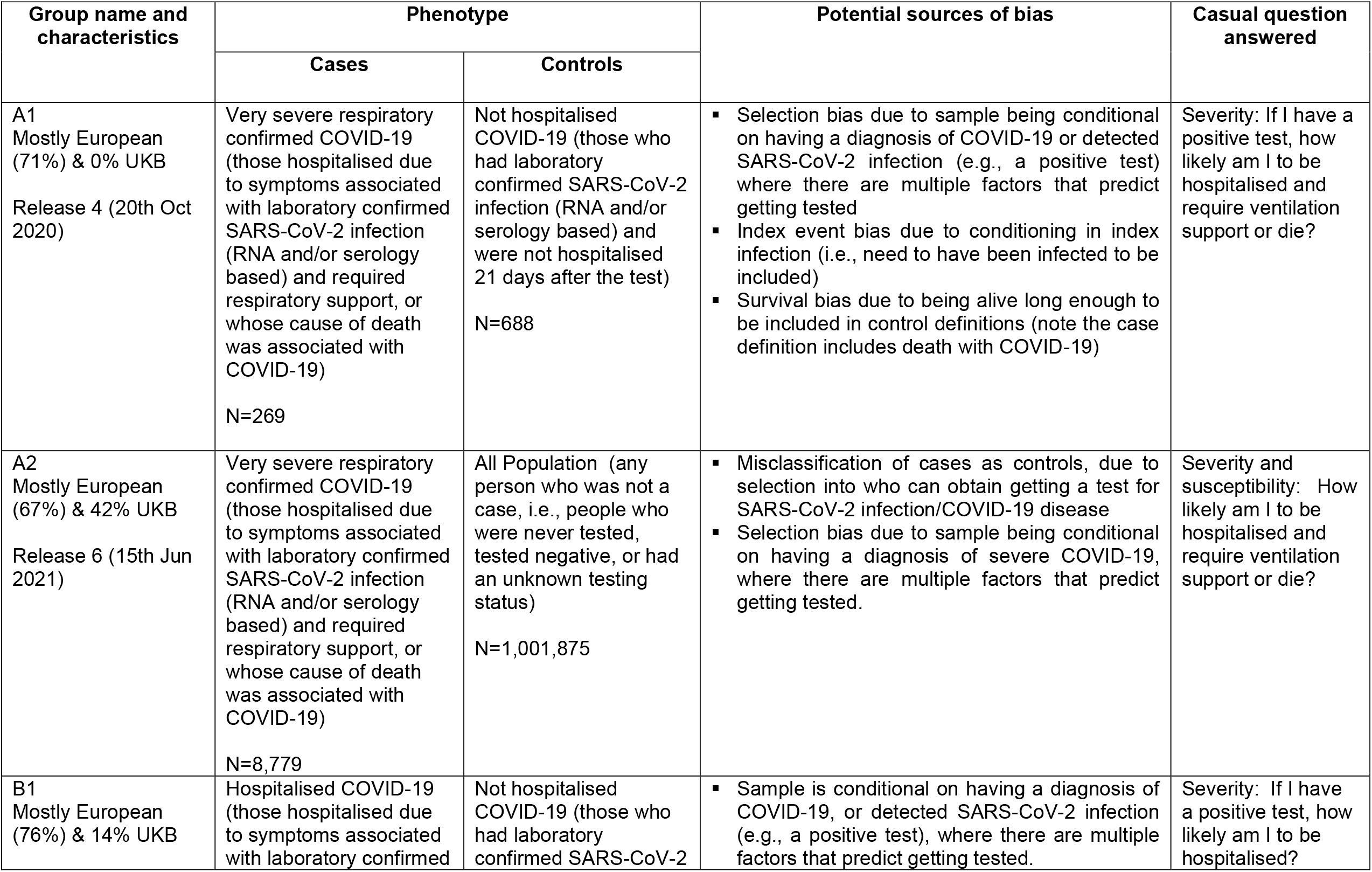
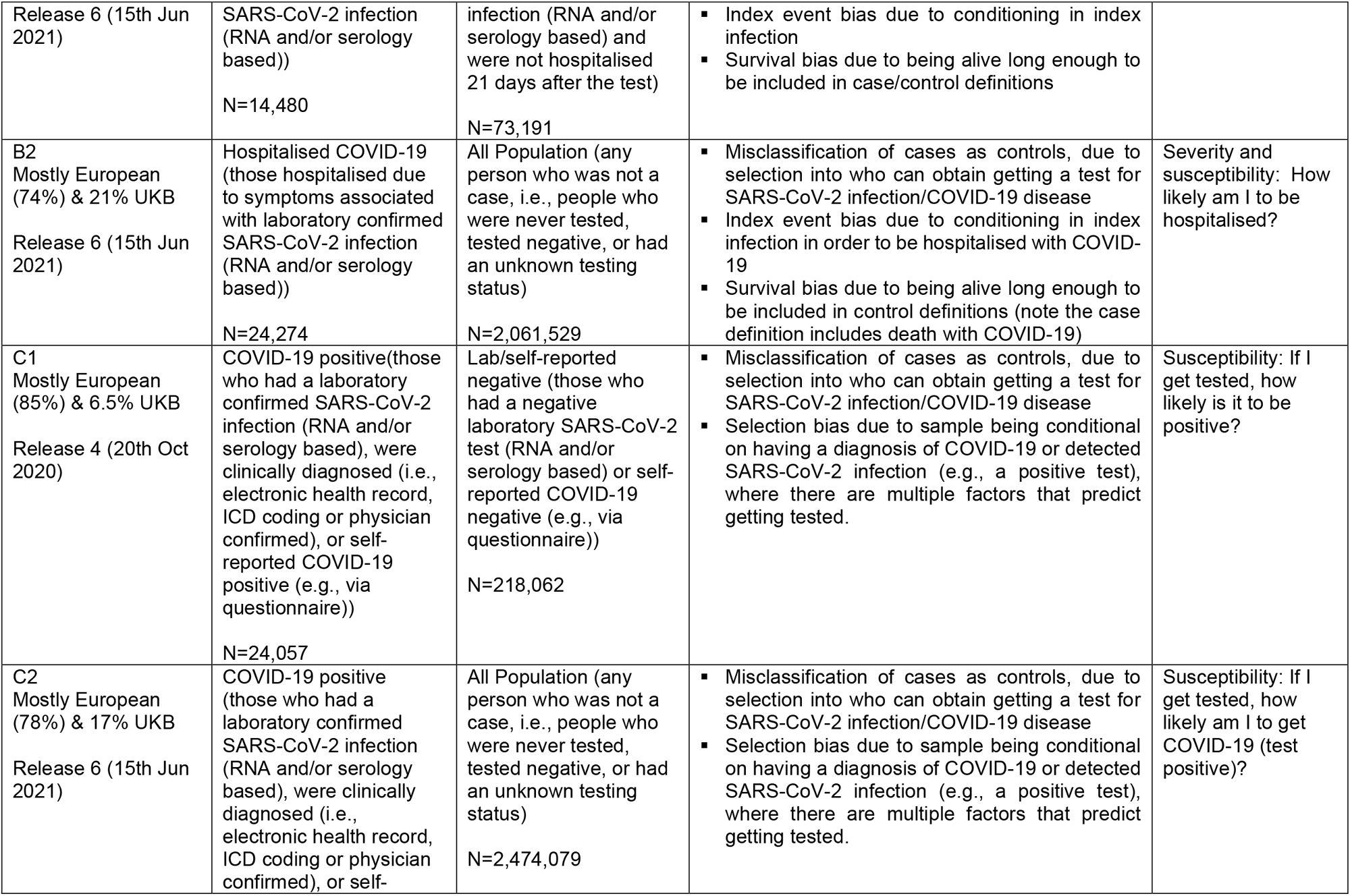
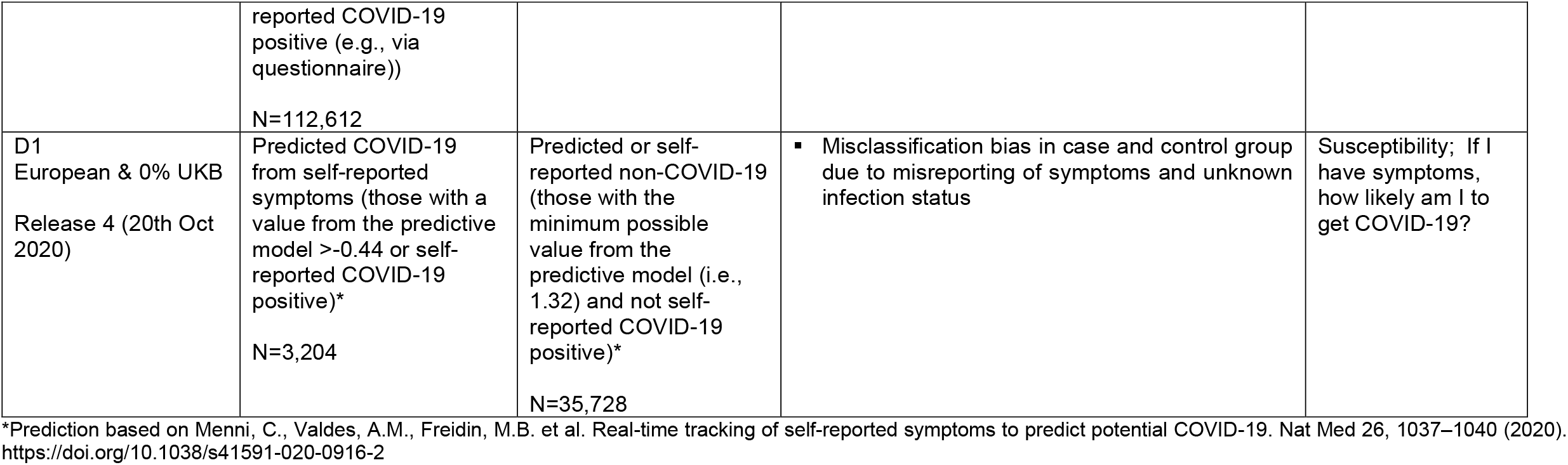
Case and control definitions (from COVID-19 Host Genetics Initiative) and causal questions.

### Misclassification bias

Misclassification happens when a study participant is incorrectly categorised for a trait/group, which could be either the exposure or the outcome (10). Whilst in MR studies non-differential measurement error can lead to weak instrument bias in the exposure and imprecision in the outcome, differential measurement error can result in bias in the observed estimates (10). Bias from misclassification of the outcome can occur when cases are wrongly classified as controls (or vice-versa). This is particularly likely to be an issue in GWAS of COVID-19 susceptibility/severity which included participants at a time when universal testing for SARS-CoV-2 infection was not available. In the UK, for example, at the start of the COVID-19 pandemic, people did not receive a SARS-CoV-2 test unless they were symptomatic or working in front-line services. As such, comparing test-positive COVID-19 participants to population controls (i.e. any person who was not a case, such as those who tested negative or whose exposure status to SARS-CoV-2 infection was unknown) could result in the controls including individuals who were infected but were asymptomatic and/or had non-severe symptoms. This would lead to misclassification bias and result in a violation of the MR exclusion-restriction criteria (11) (See **Table 1** for more details).

There have been multiple MR studies investigating the effect of body mass index (BMI) on COVID-19 susceptibility/severity (17-23), mostly concluding a causal effect of genetically predicted higher BMI on increased risk of COVID-19 susceptibility/severity. These studies are summarised in **Supplementary Table 1**. Most studies used population controls, and other than being described as a limitation, very few attempts have been made to explore selection and misclassification bias, the focus of this paper.

Several methods could be used to assess the potential influence of selection and misclassification bias, such as comparing results across different case/control comparison groups and ‘no-relevance’ control analyses. In the latter, MR analyses are carried out in a sub-population of the ‘real/relevant’ study population, in which the genetic instrument is not expected to relate to the risk factor of interest. This approach has been previously used in MR studies to assess bias due to horizontal pleiotropy (24-26). For example, in those who have never smoked, genetic variants that relate to cigarette smoking intensity would not be expected to relate to an outcome (e.g. cardiovascular disease) other than via bias, such as unbalanced horizontal pleiotropy. Thus, an interaction between variants related to smoking intensity and ever smoking can be used to determine causality, as demonstrated in a study of the potential effect of maternal gestational smoking on fetal growth parameters (26). Here, a causal effect of smoking on fetal growth is supported by seeing effects only (or stronger effects) in those who have ever smoked cigarettes, whereas seeing similar associations in those who have never smoked would be interpreted as evidence that an apparent MR causal effect is spurious. Similarly, ‘no-relevance’ studies, a form of negative control analyses, could also be used to explore selection bias. COVID-19 is a new condition that, to our knowledge, did not exist in humans before late 2019. We therefore hypothesised that any association between COVID-19, assessed in 2020 and 2021, and genetically instrumented BMI, assessed before 2019, was implausible and could only be due to bias.

In this study, we use the example of the association of BMI on COVID-19 susceptibility/severity to demonstrate methods to explore selection and misclassification bias in two-sample MR studies of risk factors for COVID-19.

## Methods

This study was reported following the recommendations of the STROBE-MR guidelines (27) (**Supplementary material**).

### Data

#### Summary genome-wide data for BMI

Summary data from the 2018 Genetic Investigation of ANthropometric Traits (GIANT) Consortium GWAS were used to genetically instrument BMI (28). This GWAS included 681,275 individuals of European ancestry, including UK Biobank participants. Sensitivity analyses were carried out using data from the previous 2015 GIANT GWAS, including data from 339,224 European-descent individuals but not from UK Biobank (29).

#### Summary genome-wide data for COVID-19

Summary data on the association of SNPs for COVID-19 susceptibility/severity were obtained from the COVID-19 Host Genetics Initiative (COVID-19 HGI) (30, 31). This initiative includes 162 contributing studies from across the world, predominantly Europe and the United States. In brief, the case/control comparisons used were:

– very severe respiratory confirmed COVID-19 vs not hospitalised COVID-19 (A1);
– very severe respiratory confirmed COVID-19 vs population, i.e. individuals from population-based cohorts, including individuals whose exposure status to SARS-CoV-2 infection was unknown or was negative (A2);
– hospitalised COVID-19 vs not hospitalised COVID-19 (B1);
– hospitalised COVID-19 vs population (B2);
– COVID-19 vs lab/self-reported negative (C1);
– COVID-19 vs population (C2);
– predicted COVID-19 from self-reported symptoms vs predicted or self-reported non COVID-19 (D1).

The definition of cases and controls for each group is detailed in **Table 1**, and the studies and number of individuals contributing to each case/control group GWAS is presented in **Supplementary Table 2**. To note, even though some groups have similar case/control definitions, different underlying studies might have contributed to different case/control comparison groups. Sample sizes varied from 957 (A1) to 2,586,691 (C2) individuals (**Table 1, Supplementary Table 2**). We used the latest data from COVID-19 HGI (release 6, from 15th June 2021) for main analyses. However, not all outcomes have been included in this release and we used the next most recent data for these (groups A1, C1 and D1). Although in October 2020 COVID-19 HGI recommended that groups A1, C1 and D1 should not be considered (no further explanations are given on the website), we used these groups in this study as the aim was to assess selection and misclassification bias, of which the different case/control definitions can be informative.

UK Biobank contributed significantly to the number of participants included in some COVID-19 GWAS (e.g., 42% of the participants for A2) and it also contributed to the GWAS we used for our main two-sample MR analyses. We undertook sensitivity analyses using a previous GWAS of BMI that did not include UK Biobank (see below under statistical analyses).

#### Selection of instruments

GWAS-significant SNPs (p<5×10^−8^) were selected as candidate instrumental variables for BMI. Proxy SNPs were used for groups A1, C1 and D1. Where a BMI SNP-outcome association was not available on the COVID-19 GWAS we sought to replace it with a proxy that was within linkage disequilibrium (LD) (and carried out in MR-Base (32)) which included 1 proxy SNP each for A1, C1 and D1 (**Supplementary Figure 2**)). LD clumping was performed based on r^2^ >0.001 using the 1000 Genomes European ancestry reference panel. Up to 507 independent SNPs for BMI were used.

For the ‘no-relevance’ study, GWAS-significant SNPs (p<5×10^−8^) were selected as candidate instrumental variables for COVID-19, and proxy SNPs were used (as defined above). For groups A1, C1 and D1, a threshold of p<5×10^−6^ was used for selection of SNPs due to one or fewer SNPs associated at conventional GWAS-significant level. LD clumping was performed as described above.

#### Data harmonisation

To ensure harmonisation, we aligned each genetic association for exposure and outcome on the same effect allele. Where possible, we used effect allele frequency (EAF) to ensure palindromic SNPs were correctly aligned.

### Main analyses

We used inverse variance weighting (IVW) with multiplicative random effects to estimate the effect of BMI on COVID-19 susceptibility/severity (33). Standard errors (SE) were corrected to account for any between-SNP heterogeneity. This method assumes that there is no directional horizontal pleiotropy. See **Figure 1 and Supplementary Table 3, and Supplementary Figure 1** for a description of each method used and MR assumptions, respectively.

**Figure 1.**
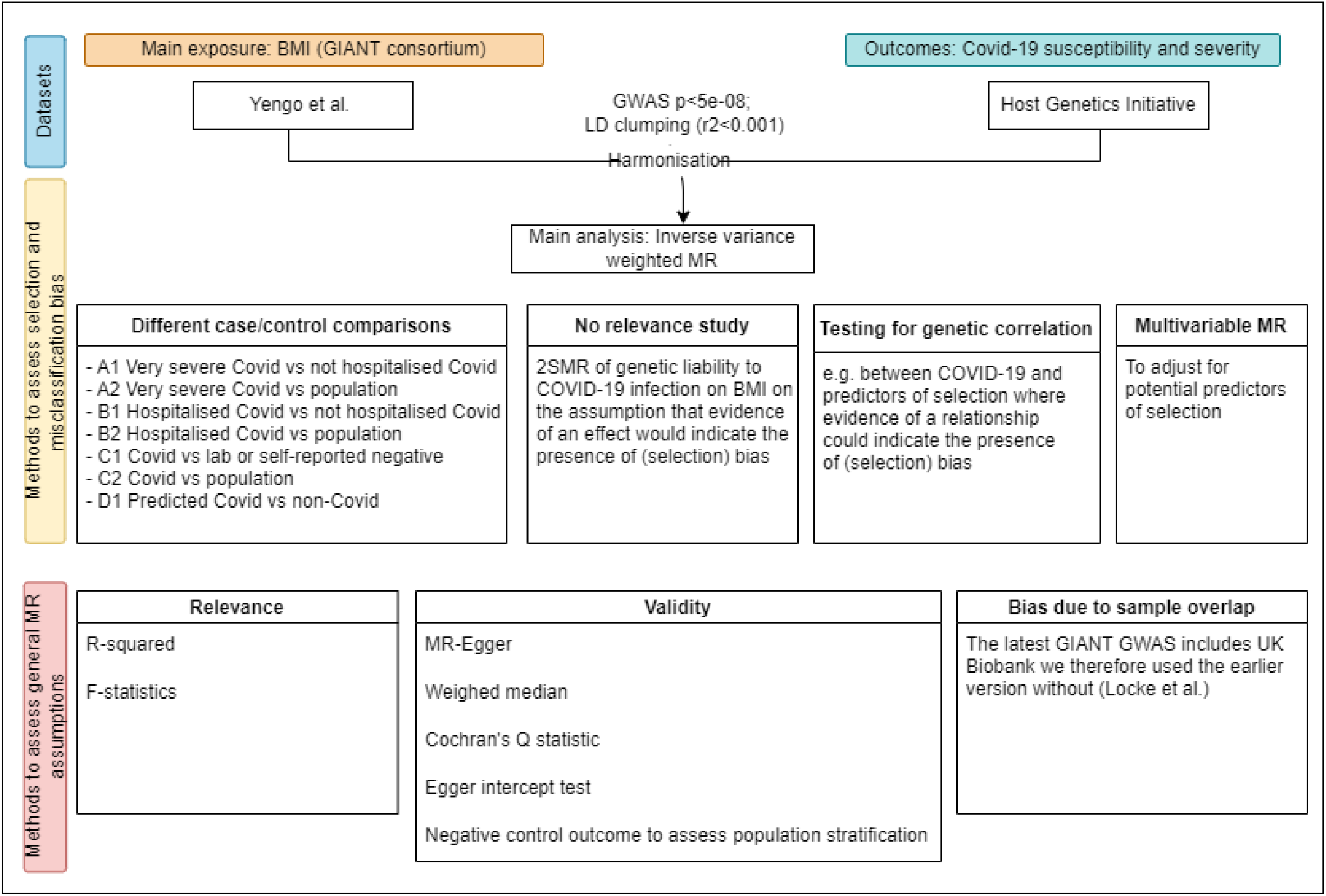
Flow chart of methods carried out to investigate selection and misclassification bias.

#### Sensitivity analyses and methods to explore the plausibility of MR assumptions

We conducted a series of sensitivity analyses to assess the validity of the core MR assumptions (5). To check the relevance assumption and weak instrument bias, we estimated the mean F statistics and total R^2^ for the combined association of all genetic variants for BMI, based on the main analysis using the most recent GIANT GWAS (i.e. including UK Biobank) for each case/control group (34, 35). For the ‘no-relevance’ analysis, we calculated the pseudo F-statistic and R^2^ both with (36) and without (37) assuming a prevalence for each case control comparison based on external information (38). We checked for between-SNP heterogeneity in the main IVW analyses using the Cochran’s Q test and undertook sensitivity analyses using MR-Egger (33) and weighted-median (39), which, when certain conditions are met, would be unbiased in the presence of unbalanced horizontal pleiotropy. Single-SNP analysis and leave-one-out analysis were also conducted. As UK Biobank contributed to both the COVID-19 HGI and the most recent GIANT GWAS for BMI, we also repeated analyses (main analysis only) using summary results from a previous GIANT BMI GWAS, which does not include UK Biobank, as a sensitivity analysis to explore any bias due to sample overlap. Analyses were repeated using 79 independent SNPs and results were compared to our main analysis results.

To assess bias caused by population stratification, i.e. confounding by population (see **Table 2**), we used skin tanning as a negative control outcome to compare the association observed between BMI and COVID-19 with the association observed between BMI and skin tanning (7). Here, a negative control would be a variable that is not expected to be associated with the outcome but has the same underlying population structure. Evidence of an association when using the negative control outcome could indicate bias from population stratification. For this analysis, we also used the BMI GWAS not including UK Biobank. We similarly explored bias in the COVID-19 GWAS by using COVID-19 as the exposure and skin tanning as the outcome.

**Table 2.** Summary of Mendelian randomisation methods carried out to investigate selection bias and misclassification bias in analyses of body mass index (BMI) and COVID-19 susceptibility and severity

| Method | Description |
| --- | --- |
| Different case-control definitions | Whilst Mendelian randomisation (MR) is said to be unbiased due to measurement error, genome-wide association studies (GWAS) can be biased due to an incorrectly specified phenotype, such as by misclassification of the phenotype. Additionally, selection into who is included in the GWAS may introduce bias. In the context of COVID-19, few of the populations contributing to GWAS had universal testing for infection, and participants with COVID-19 may have been incorrectly classified as controls. There could also be systematic differences between cases and controls related to factors influencing one's probability of being assessed, introducing selection bias.<br>Examining the consistency of effects across different cases and control definitions allows for assessment of how different sources of misclassification bias or selection bias may introduce error. Where results are consistent across definitions (e.g., COVID-19/population controls vs COVID-19/ lab or self-report negative), the evidence for a causal effect is greater. However, where effect estimates diverge across different definitions, this may be evidence to suggest selection bias or misclassification bias is present. |
| 'No-relevance' study | No-relevance studies have previously been used in MR analyses to test for pleiotropy (24). Here, using a similar principle, we use it to test for selection bias. MR analyses are carried out in a sub-population or the same population where the genetic instrument is not expected to relate to the risk factor of interest, but is subject to the same selection mechanism. In this case, where the outcome (BMI) has been measured before the exposure (COVID-19 susceptibility and severity) existed. Furthermore, this is similar to a negative control design, where any evidence of an association is likely due to bias, rather than a true causal effect. |
| Testing for genetic correlation between SNPs | SNPs for different phenotypes should be uncorrelated. Testing for overlapping SNPs between COVID-19 (or indeed any other exposure/outcome) and hypothesised predictors of selection identifies whether a GWAS may be associated with the phenotype of interest, or whether the GWAS is identifying factors associated with a diagnosis. In this case, that would be factors associated with obtaining a test for SARS-CoV-2, i.e. smoking, income and education. |
| Multivariable Mendelian randomisation | Multivariable MR (MVMR) can estimate the causal direct effects of multiple exposures, particularly to assess whether SNPs for an exposure may exert a pleiotropic effect on a second variable, or to test for mediators. MVMR can also offer a method to adjust for potential selection. This approach has previously been described in the non-instrumental variable literature as a method to account for selection bias (48). A direct effect, estimated via MVMR, attenuated from the unadjusted total effect can indicate any one of pleiotropy, mediation, or selection bias. Therefore, researchers would need to use subject matter knowledge to determine which would be most likely and carefully consider what variables may be considered predictors of selection in an analysis. |
| Negative control exposure/outcome | We have used a negative control study to test for population stratification. However, it is also possible to use a negative control design to test for selection bias. Here, a negative control would be a variable that is not expected to be associated with the exposure/outcome, but is subject to the same selection mechanisms. Any evidence of an association would therefore indicate selection bias is present. In this analysis, whilst many variables have been shown to be associated with selection, they have also been shown to be associated with BMI and/or COVID-19 susceptibility/severity, and therefore would not satisfy the conditions of being a negative control (18). |

Note that these methods are not explicitly for testing for selection bias and can be used to test for different sources of bias in Mendelian randomisation.

### Methods to explore potential misclassification and selection bias

We compared different case/control definitions to explore potential misclassification or selection bias. For example, where the case definition remains the same, e.g. for C1 (COVID-19 positive vs lab/self-report negative) and C2 (COVID-19 positive vs population), only those tested for SARS-CoV-2 (or those who self-reported COVID-19 status) could be included as cases, and selection bias can be present due to individual characteristics related to having a test or self-reporting COVID-19 status, and therefore being in the analysis. However, when we compare COVID-19 positive cases to population controls (C2), then we make the assumption that those who have not been tested are negative (hence classified as controls), i.e. differential misclassification due to individual characteristics being related to getting tested, and therefore being misclassified. As such, if effect estimates are similar across different control definitions, then it suggests selection or misclassification bias may not be present. If the point estimates differ, this may indicate selection or misclassification bias is present. More details are given in **Table 1** and **Table 2**.

We used COVID-19 as the exposure and BMI as the outcome in a ‘no-relevance study’ to determine whether the genetic instruments for COVID-19 were related to BMI. Given COVID-19 could not influence BMI assessed prior to 2019 (i.e., before COVID-19 existed), plausibly we would expect null findings. If any effects of COVID-19 on BMI are observed, this suggests the presence of selection bias, or other biases such as horizontal pleiotropy, and indicates that effects of BMI on COVID-19 (main analysis) may be similarly biased.

It is possible that some of the COVID-19 SNPs are identifying factors associated with getting tested rather than COVID-19 *per se* (40). For example, we know that being a non-smoker and being more highly educated are predictors of getting a test (14). Using linkage disequilibrium score regression (LDSC), we estimated the genetic correlation between COVID-19 SNPs and SNPs associated with potential predictors of testing (smoking, education and income) (40). These potential predictors of selection were determined *a priori* from estimates in the previous literature (13, 14, 41) and due to the availability of GWAS summary statistics.

We then used multivariable MR (MVMR) to adjust for these predictors of being tested to estimate a direct effect of BMI on COVID-19 independent of potential influences on being tested (42). Evidence of a direct effect which is different to the total effect in the main IVW would support the presence of selection bias.

To identify whether the effects of misclassification and selection bias may have changed over the course of the pandemic (given more studies and more participants were included in more recent GWAS), we repeated the main and the ‘no-relevance’ analyses for two of the comparison groups. We used hospitalised COVID-19 vs not hospitalised COVID-19 (B1), and COVID-19 vs population (C2) using different releases of the GWAS data covering time periods up to July 2020 (release 3), October 2020 (release 4), January 2021 (release 5) and June 2021 (release 6). For the other groups, repeat releases of GWAS summary data across all time points were not available.

#### Software and code

All analyses were conducted using R version 4.1.1. Two-sample MR analyses were performed using the “TwoSampleMR” package version 0.5.5 available at https://mrcieu.github.io/TwoSampleMR/. Multivariable MR analyses were conducted using the “MVMR” package version 0.3 available at https://github.com/WSpiller/MVMR. LDSC was conducted using the LDSC GitHub repository available at https://https://github.com/bulik/ldsc. Code for all analyses is available at https://github.com/gc13313/BMI_COVID_2SMR.

### Patient and public involvement

Neither patients nor members of the public were involved in the original study design. The idea and subsequent results were presented to meetings of the British Heart Foundation / National Institute of Health Research COVID Highlight projects (COVIDITY-COHORT group), with feedback on ways of explaining the concepts to diverse audiences and readers.

## Results

### Illustrative example: Two-sample MR of potential effect of BMI on COVID-19 in different case/control groups

From the 507 SNPs independently associated with BMI, after data harmonisation the number of SNPs ranged from 487 (group A2) to 490 (group D1) (**Supplementary Figure 2**). The mean F-statistic for the combined association of these SNPs for BMI was 72.8, explaining 5% of the variation in BMI.

In IVW estimates, we found that genetically higher BMI was associated with higher odds of COVID-19 for all groups assessed, except A1 (very severe COVID-19 vs not hospitalised COVID-19, OR 0.56 per 1 SD higher BMI, 95%CI 0.22, 1.41) which had the smallest sample size (N_cases_ = 269; N_controls_ = 688). For the remainder, odds ratios ranged from 1.14 (95%CI 1.11, 1.18) for group C2 (COVID-19 vs population) to 1.57 (95%CI 1.39, 1.78) for group A2 (very severe COVID-19 vs population) (**Figure 2(a)**). Results were consistent when the same case outcome was compared to different control groups (**Figure 2(a)**). Sensitivity analyses using MR-Egger and weighted-median showed consistent results to the main IVW results, though with expected wider confidence intervals (**Supplementary Figure 3(a)**). Sensitivity analyses using the previous GIANT GWAS not including UK Biobank showed similar findings, although, again, with expected wider confidence intervals (**Supplementary Figure 4**). There was some evidence of between-SNP heterogeneity for groups A2, B2 and C2 (**Supplementary Figure 3(a)** and **Supplementary Table 4)** and no strong evidence of horizontal pleiotropic effects (**Supplementary Figure 3(a)** and **Supplementary Table 5**).

**Figure 2.**
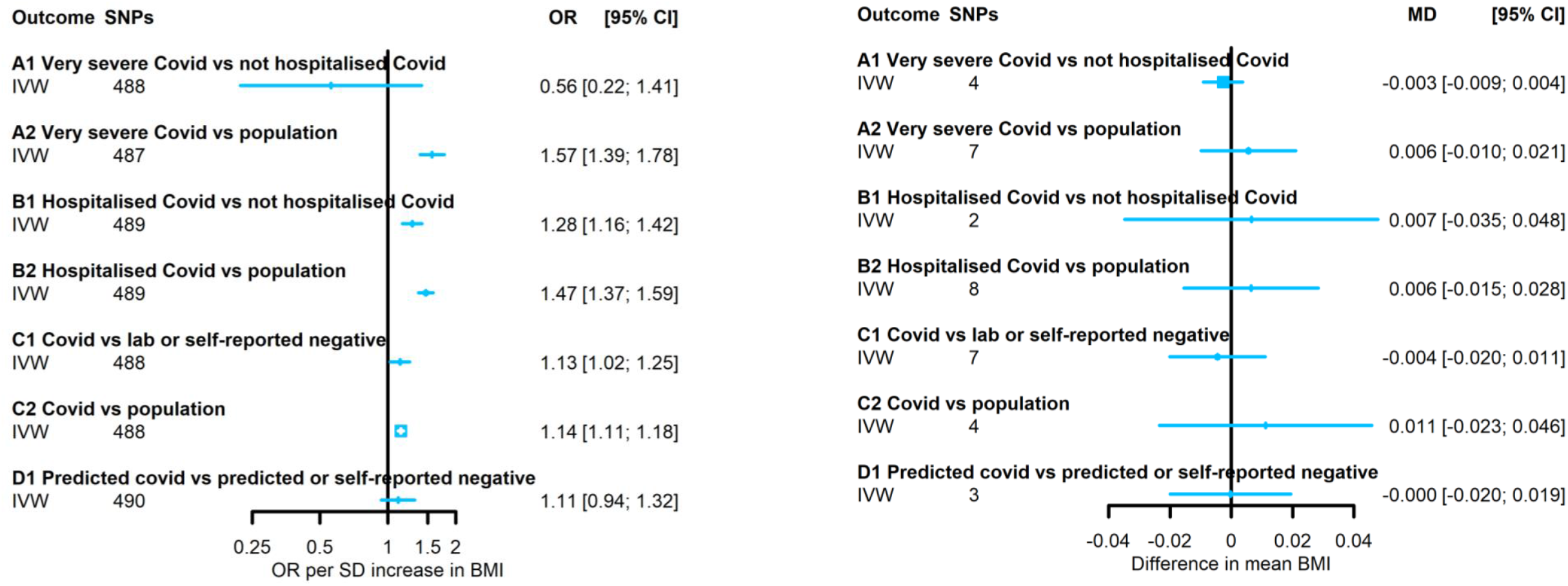
(a) Main analysis: Effect of body mass index (BMI) on SARS-CoV-2/ COVID-19 (b) ‘No-relevance analysis’: Effect of SARS-CoV-2/COVID-19 on BMI. Footnote: IVW: inverse variance weighting; MD: mean difference; OR: odds ratio; SNPs: single nuclear polymorphism

### Analysis to explore population stratification

The association between BMI and ease of skin tanning (a negative control outcome to test for population stratification) showed results close to the null (mean difference [MD] in ease of skin tanning: 0.01; 95% CI: -0.02 to 0.04 per SD increase in BMI) (**Supplementary Figure 5(a)**). The MR association between all COVID-19 case-control groups and skin tanning was also close to the null (e.g., COVID-19 vs population control (C2), MD in ease of skin tanning; -0.02; 95% CI: -0.05 to 0.02) (**Supplementary Figure 5(b)**).

### Analyses to assess whether potential misclassification and selection bias explain some of the identified effects in our illustrative analyses

#### ‘No-relevance’ analysis

The number of SNPs (including proxies) available after harmonisation varied from two (group B1) to eight (group B2), and the mean pseudo-F-statistics varied from 21.7 (group D1) to 134.2 (group C2) (**Supplementary Table 6**). The percent variation of COVID-19 explained by the combined SNPs included after harmonisation varied from 0.02% (group C2) to 10.6% (group A1).

For all seven case/control groups in the ‘no-relevance’ analyses, the effect estimates of genetic liability to COVID-19 on BMI measured pre-pandemic were all close to the null (**Figure 2(b)**), with results consistent when using MR-Egger and weighted-median methods (**Supplementary Figure 3(b)**). There was some evidence of between-SNP heterogeneity in groups A1, A2 and B2 for the associations between genetic liability to COVID-19 and BMI (**Supplementary Table 4** and **Supplementary Figure 6**) and some evidence of horizontal pleotropic effects in group A1 (**Supplementary Table 5**).

#### Genetic correlation

There was evidence from LDSC analyses of genetic correlation between most groups of COVID-19 outcomes and BMI, smoking, education, and income, except for groups C1 and D1. The correlations of COVID-19 outcomes with BMI and smoking were positive, and negative with education and income. Correlations were typically modest to weak (ranging from -0.34 for group C1 and education to 0.35 for group A2 and BMI) (**Table 3**). The genetic correlation with each predictor of getting tested (selection) was overall similar across the different case/control comparison groups.

**Table 3.** Linkage disequilibrium score regression (LDSC) of body mass index (BMI) and SARS-CoV-2/COVID-19 comparisons groups with possible predictors of selection.

|  | BMI |  | Smoking |  | Education |  | Income |  |
| --- | --- | --- | --- | --- | --- | --- | --- | --- |
|  | rG (se) | P value | rG (se) | P value | rG (se) | P value | rG (se) | P value |
| A1: Very severe COVID-19 vs not hospitalised COVID-19* | n/a |  | n/a |  | n/a |  | n/a |  |
| A2: Very severe COVID-19 vs population | 0.35 (0.05) | 1.08x10<sup>-11</sup> | 0.23 (0.08) | 0.003 | -0.24 (0.04) | 1.07x10<sup>-8</sup> | -0.27 (0.06) | 5.99x10<sup>-6</sup> |
| B1: Hospitalised COVID-19 vs not hospitalised COVID-19 | 0.30 (0.06) | 5.63x10<sup>-8</sup> | 0.19 (0.08) | 0.027 | -0.20 (0.05) | 3.11x10<sup>-5</sup> | -0.20 (0.06) | 0.002 |
| B2: Hospitalised COVID-19 vs population | 0.35 (0.04) | 1.32x10<sup>-5</sup> | 0.25 (0.06) | 6.34x10<sup>-5</sup> | -0.33 (0.04) | 5.59x10<sup>-19</sup> | -0.26 (0.05) | 7.99x10<sup>-8</sup> |
| C1: COVID-19 vs lab or self-reported negative | n/a |  | 0.13 (0.44) | 0.763 | -0.68 (0.75) | 0.364 | -0.26 (0.82) | 0.752 |
| C2: COVID-19 vs population | 0.32 (0.04) | 1.01x10<sup>-14</sup> | 0.13 (0.07) | 0.062 | -0.34 (0.04) | 2.01x10<sup>-18</sup> | -0.19 (0.05) | 7.35x10<sup>-5</sup> |
| D1: Predicted COVID-19 vs predicted or self-reported negative | 0.10 (0.09) | 0.268 | 0.20 (0.21) | 0.334 | -0.02 (0.07) | 0.757 | -0.19 (0.11) | 0.070 |
| BMI | - |  | 0.28 (0.04) | 7.09x10<sup>-12</sup> | -0.27 (0.01) | 6.48x10<sup>-76</sup> | -0.22 (0.02) | 1.20x10<sup>-33</sup> |

rG: Genetic correlation between two traits; se: standard error
\*Small heritability for this phenotype and therefore genetic correlation cannot be estimated

#### MVMR adjusting for potential predictors of selection

Results from MVMR adjusted for smoking, education and income were similar to the main analysis (**Figure 3**).

**Figure 3.**
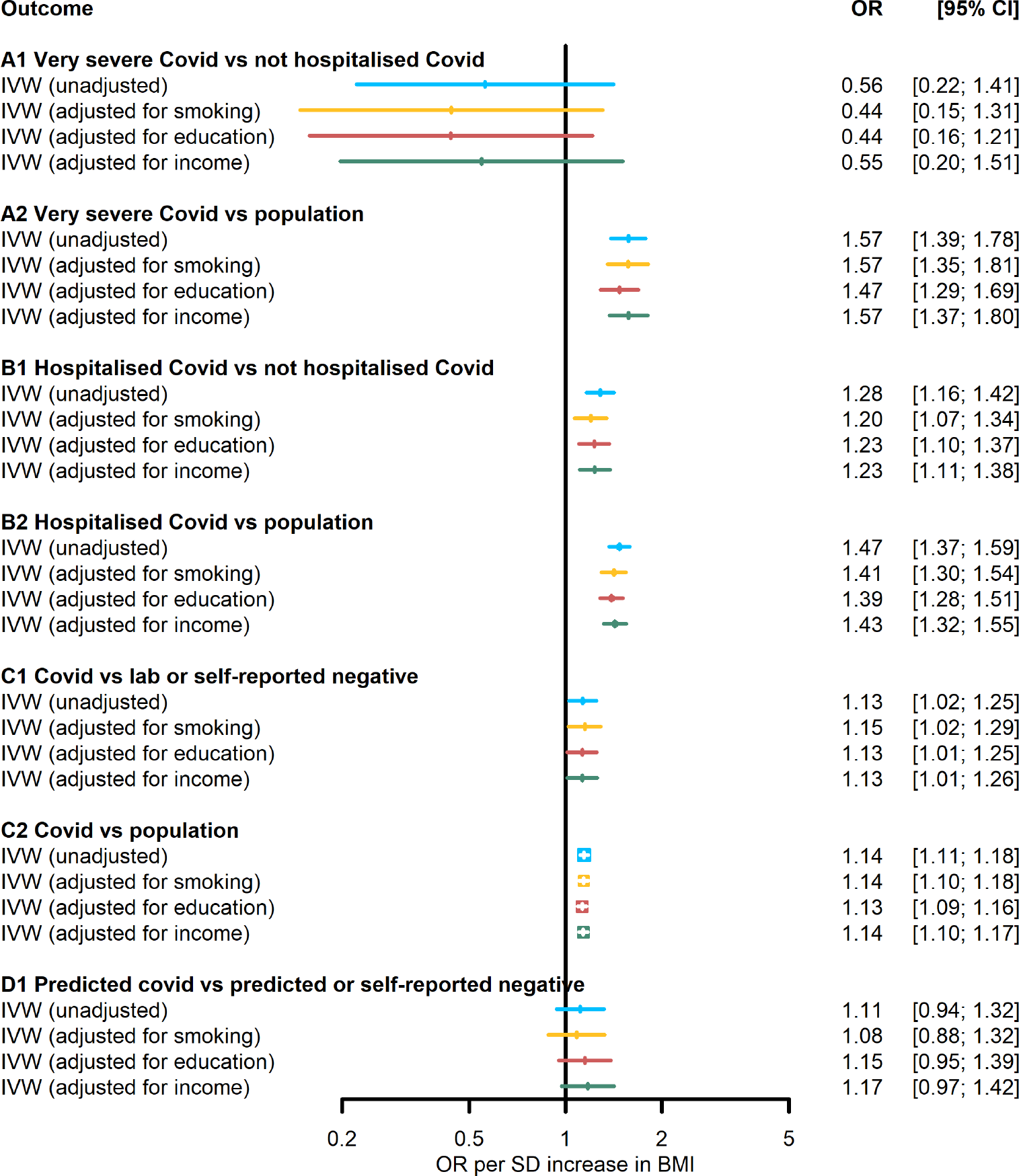
Multivariable Mendelian randomisation (MVMR): Effect of body mass index (BMI) on SARS-CoV-2/COVID-19 adjusted for education, smoking and income. Note: Education (years of schooling, ieu-a-1239); smoking (cigarettes per day, ieu-b-25) and income (ukb-b-7408

#### Comparisons over time

The point estimate for the OR between BMI and COVID-19 severity (group B1) increased over the course of the pandemic (OR: 0.89; 95% CI: 0.51 to 1.53 based on data from release 3 compared with OR: 1.28; 95% CI: 1.16 to 1.42 based on data from release 6) (**Figure 4(a)**), although confidence intervals between the timepoints overlapped, reflecting differences in power and precision. Meanwhile, the point estimate for the OR between BMI and COVID-19 susceptibility (group C2) decreased over time (OR: 1.41; 95% CI: 1.21 to 1.63 based on data from release 3 compared with OR: 1.14; 95% CI: 1.11 to 1.18 based on data from release 6). With the exception of the negative association between COVID-19 severity (as the exposure) and BMI based on COVID-19 data collected up to July 2020 (mean difference: -0.010, 95% CI -0.016, -0.003), the ‘no relevance’ analyses were consistent with the null throughout all data releases (**Figure 4(b)**). As expected, since sample sizes for the GWAS increased, all results became more precisely estimated over time.

**Figure 4.**
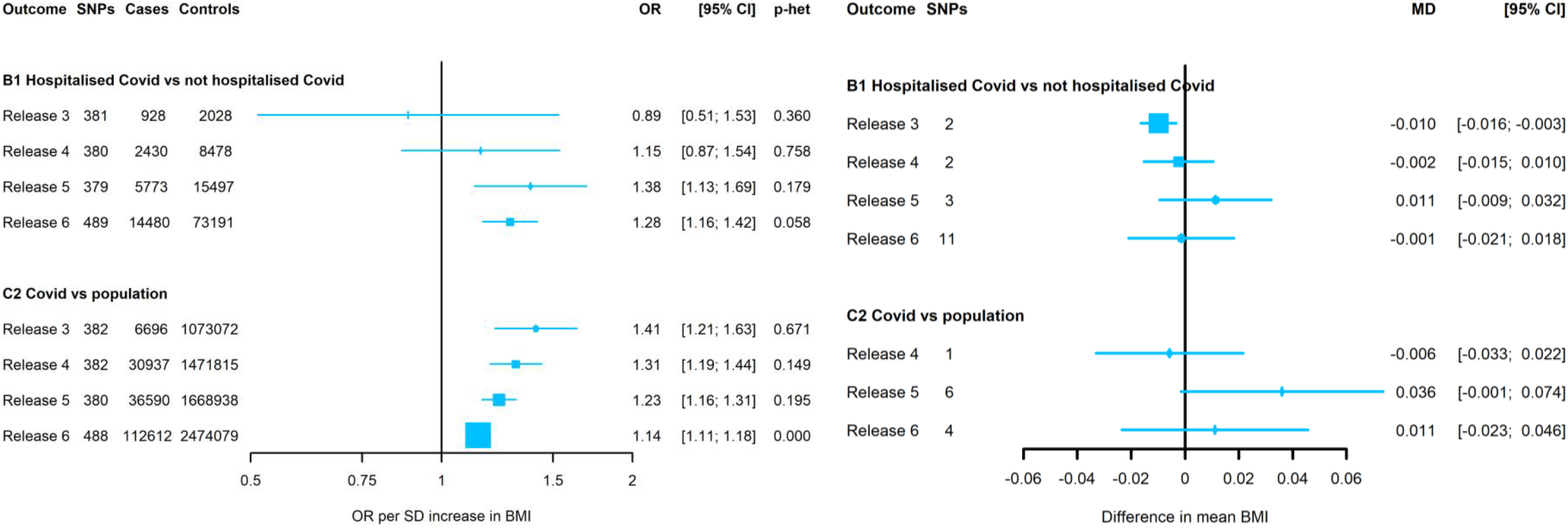
(a) Main analysis: Effect of body mass index (BMI) on SARS-CoV-2/COVID-19 and (b) Effect of SARS-CoV-2/COVID-19 on BMI - by release of COVID-19 Host Genetic Initiative release date. Footnote: OR: odds ratios; SNPs: single nuclear polymorphism. OR correspond to inverse variance weighing (IVW) estimates. Release 3: released on 2^nd^ July 2020; release 4: released on 20^th^ October 2020; release 5: released on 18^th^ January 2021; release 6: released on 15^th^ June 2021. For comparison group B1 a threshold of p<5×10^−6^ was used for selection of SNPs (across all releases to allow for comparison) due to the inexistence or small number of SNPs associated at conventional GWAS-significant level. No SNPs were available for C2 at release 3. Note that these are not mutually exclusive samples over time and different SNPs predict COVID-19 each time.

## Discussion

### Principal findings

In this two-sample MR analysis, we have identified evidence to suggest that BMI is a risk factor for COVID-19 susceptibility/severity. Using a range of sensitivity analyses, including a ‘no-relevance’ analysis and MVMR to adjust for variables associated with selection, we have demonstrated that selection into COVID-19 GWAS is likely, but that this does not appear to result in substantial bias in MR analyses of BMI on COVID-19 susceptibility/disease. Neither do we find evidence for misclassification bias. This should not be interpreted as reassurance for <u>all</u> MR studies of potential risk factors for COVID-19, as the effects of any selection and/or misclassification will differ between different exposures. The methods we have used here (and for which we provide complete code) would be useful to undertake in any MR studies of COVID-19 and for other causal questions using MR where there are concerns about selection bias.

#### Strengths and limitations of the study

To our knowledge this is the first MR study of COVID-19 outcomes to take a systematic approach to explore selection and misclassification bias in this area, including a novel application of ‘no-relevance’ analyses and exploiting time-varying associations. In our illustrative example and in the ‘no-relevance’ analyses we have conducted appropriate sensitivity analyses to assess the plausibility of the MR assumptions, including comparing estimates from a BMI GWAS without UK Biobank participants included and a negative control design to explore possible bias due to population stratification (7).

The summary statistics used for COVID-19 are from the largest available GWAS, incorporating many global studies with differing selection processes (31). The ‘no-relevance’ analysis results may have been biased by weak instruments, as there were far fewer SNPs for any COVID-19 instruments than for BMI (the exposure in our ‘real’ illustrative example). Weak instrument bias in two-sample MR is expected to bias estimates towards the null. However, the F-statistics for the ‘no-relevance’ COVID-19 instruments varied from 22 to 134, suggesting weak instrument bias is unlikely. For some of the COVID-19 exposures in the ‘no-relevance’ analyses we had to relax the p-value threshold in order to have genetic instruments, and this could result in increased likelihood of horizontal pleiotropy. Consistent results from MR-Egger, weighted median analyses, and the ‘no-relevance’ analyses, including analysis over time, where sample sizes increased, suggest that any weak instrument or horizontal pleiotropy bias is likely to have been small.

In our illustrative example, all our outcomes were binary and we have carried out analyses on the log odds ratio scale in order to compare results of the effect of BMI on COVID-19 with others that have been published (17-23). The symmetrical nature of the odds ratio, where an interaction between the exposure (BMI) and the outcome (COVID-19) is on the multiplicative scale, means that the exposure and outcome would need to be associated with selection for the odds ratio to be biased (43). As we have carried out analyses on the log odds ratio scale, as opposed to the risk difference scale, the magnitude of any biases here may be smaller than those observed for a continuous outcome, or for analyses of COVID-19 carried out on the risk difference scale.

Our ‘no-relevance’ analyses is valid if our assumption that liability to COVID-19, which did not exist before late 2019/early 2020, could not influence outcomes such as BMI, that already existed (were assessed) before. We feel this is plausible but, as with other ‘no-relevance’ analyses, violation of this assumption cannot be ruled out. For example, susceptibility to COVID-19 is likely influenced by innate immunity and may also be influenced by acquired immunity from other infections. Thus, in other MR studies if an association in the ‘no-relevance study’ is similar to that of the ‘real analyses’, interpreting that as bias in the main analysis might not be correct. This is the reason why we feel that exploring bias through a number of approaches, including exploring factors related to selection, undertaking multivariable MR with those, and comparing different case/control groups is important, as well as considering for specific questions the extent to which the ‘no-relevance’ assumption might be violated. Indeed, previous analyses exploring the association between COVID-19 on stroke have been framed as such (44).

We saw consistent results across all comparisons except for A1. This may be because this comparison group is at the most risk of selection bias as ‘very severe respiratory confirmed COVID-19’ depends on getting a test and depends on being hospitalised. It was also one of the comparisons which the COVID-19 HGI said should not be considered, although no reason was given. However, results adjusting for predictors of selection showed similar results to the main analysis.

#### Comparison to other studies

Previous studies that have highlighted and/or explored the potential impact of selection bias in MR with other outcomes (16) have been mainly focused around one sample MR, and reweighting based on the target population via inverse probability weighting (IPW) (6) and multiple imputation, where applicable (45). However in a two-sample MR setting, multivariable MR adjusting for factors associated with selection and the outcome (46) and the use of negative control exposures (11) have also been recommended.

In relation to our illustrative example, a number of studies (summarised in **Supplementary Table 1**) have sought to assess the potential causal effect of BMI on COVID-19 susceptibility and severity using two-sample MR with the COVID-19 HGI GWAS (17-23). Whilst misclassification and selection bias are often raised as study limitations, previous MR studies of BMI and COVID-19 have not explored the impact of these biases on the robustness of the results. Although some studies have considered different case/control comparison groups as different outcomes (17), these have typically not considered that each group may be subject to different biases or how the different groups may inform identifying the presence of potential bias.

Previous studies using summary data from the COVID-19 HGI have used releases three (18), four (17, 22, 23) and five (21) (see Table S1 in supplementary material). In comparison to analyses from release 4, effect estimates for the association between BMI and COVID-19 susceptibility and severity in our analyses using release 6 were smaller and estimated with greater precision, potentially reflecting larger samples and greater power. As testing capacity increases globally, there may be less misclassification of population controls in the more recently included data, and less selection bias in who can obtain a test for SARS-CoV-2 infection. Alternatively, as the pandemic persists and behaviours change, risk factor estimates may change over time. Other changes, such as vaccination (and, for example, that BMI could have a differential effect on how vaccination influences immunity (47)) could also influence estimates. This demonstrates the importance in continuing to update risk factor estimates as new data are released and the pandemic continues, to ensure that public health approaches are optimally used to combat risk of COVID-19. If risk factors change over time, then public health approaches may also need to adapt.

#### Meaning of the study

The aim of this study was to explore methods for identifying the effects of selection and misclassification bias in MR analyses that seek to identify risk factors for different COVID-19 outcomes. This is important as the need to rapidly understand key determinants of these outcomes as the pandemic has, and continues to unfold, whilst balancing that with the need for unbiased/minimally bias results is paramount. Risk of selection bias in this context is high, given, for example, pressures on who does and does not experience and/or report symptoms, who gets tested (whether symptomatic or not) and how that has changed over time. Our *a priori* assumption was that genetic, including GWAS and MR, analyses are prone to bias from such selection. We are not aware of any previous studies that have explicitly explored such bias in MR studies of COVID-19 outcomes.

Given that selection and misclassification bias are an issue in this area, we have systematically carried out a range of methods, as outlined in **Table 2**, to help test for these biases using an illustrative example of BMI and COVID-19. The impact of selection bias and misclassification will depend on the variables included in an analysis and on the underlying data used. Therefore, although we did not find evidence of bias in our illustrative example of BMI and COVID-19, we recommend that researchers consider potential misclassification and selection bias in MR analyses, particularly when using highly selected phenotypes or disease outcomes. We suggest researchers to look across the different methods used here and consider whether together they suggest no selection, misclassification bias or some other sources of bias. However, depending on the available data, it may not be possible to conduct every sensitivity analysis.

#### Unanswered questions and future research

As summary statistics for COVID-19 continue to be updated, these analyses should be used with the methods we suggest (including the time variation assessment) to improve understanding of the aetiology of COVID-19 susceptibility and severity. Future effect estimates obtained using summary data may be less prone to selection bias as universal testing becomes more widespread, particularly in most high-income countries, and salient symptoms have been defined for adults. However, some selection is likely to remain, such as if those whose jobs and livelihood depend on being tested negative either do not get tested or do not report positive results.

As individual studies with genotypic data obtain more data on COVID-19 in their study samples, it may be possible to have enough power to carry out participant-level MR analyses. Where individual-level data are available, further methods are available for exploring and/or accounting for selection bias, including IPW, which can be used to weight the study sample to better reflect the population of interest (6). Index event bias may be present in MR analyses of COVID-19 disease progression (48) which can occur when conditioning on an index event (i.e., SARS-CoV-2 infection). Future work to account for this bias in MR analyses is required.

#### Conclusion

We have proposed a set of analyses for exploring potential selection and misclassification bias in MR studies of potential risk factors for SARS-CoV-2 infection and COVID-19. We have demonstrated the use of these with an illustrative example of the effect of BMI on COVID-19 outcomes. Although these found presence of selection by socioeconomic position and related traits, for this particular example, the MR results are not substantially affected by selection/misclassification bias. We recommend that the methods we demonstrate and provide detailed analytic code for are used in MR studies of risk factors for COVID-19 and other MR studies where such biases are likely in the available data.

## Acknowledgments

We are extremely grateful to the British Heart Foundation-National Institute of Health Research (NIHR) COVIDITY-COHORT group for their helpful discussions throughout. We also thank the COVID-19 HGI and GIANT consortia for making GWAS summary statistics publicly available. The acknowledgment to the people and studies/projects that contributed data to COVID-19 HGI is available at <https://www.covid19hg.org/acknowledgements/>.

## Availability of data and materials

The datasets analysed during the current study are all publicly available GWAS. Analysis scripts and the analysis plan can be found on the following GitHub page: https://github.com/gc13313/BMI_COVID_2SMR.

## Ethical approval and consent to participate

Ethics approval was not required for this study, given only summary-level (GWAS) data has been used.

## Funding

This work was supported by the Bristol British Heart Foundation (BHF) Accelerator Award (AA/18/7/34219, which supports ARC); the University of Bristol and Medical Research Council (MRC) Integrative Epidemiology Unit (MC_UU_00011/1, MC_UU_00011/3, MC_UU_00011/6, which supports ARC, GLC, AGS, NG, GDS, DAL, KT, and MCB); the BHF-National Institute of Health Research (NIHR) COVIDITY flagship project; the European Union’s Horizon 2020 research and innovation programme (under grant agreement No 733206 LifeCycle, which supports GLC and DAL); the Dynamic longitudinal exposome trajectories in cardiovascular and metabolic non-communicable diseases (H2020-SC1-2019-Single-Stage-RTD, project ID 874739, which supports AGS); the University of Bristol (Vice-Chancellor’s Fellowships, which supports MCB); the British Heart foundation (CH/F/20/90003, to DAL); and the NIHR (NF-0616-10102, to DAL).

The funders had no role in study design, data collection and analysis, decision to publish, or preparation of the manuscript. For the purpose of Open Access, the author has applied a CC-BY public copyright licence to any Author Accepted Manuscript version arising from this submission.

This publication is the work of the authors and Gemma L Clayton, Ana Gonçalves Soares, Neil Goulding, Deborah Lawlor and Alice R Carter will serve as guarantors for the contents of this paper.

## Author contributions

DAL conceived the study. All authors designed the study. GLC wrote the analysis plan with input from all authors. ALGS, NG, GLC and ARC performed the analyses. GLC and ARC wrote the original draft of the manuscript with input from all authors. All authors were involved in the interpretation of results, helped refine the manuscript, and approved its final version.

