## Supplemental figures and tables for "The relationship between BMI and COVID-19: exploring misclassification and selection bias in a two-sample Mendelian randomisation study"

### Supplementary figures and tables


| |
|---|
| Supplementary Table 1 Summary of one/two sample Mendelian randomisation (MR) studies that assessed the effect of body mass index (BMI) on SARS-CoV-2/COVID-19 |
| Supplementary Table 2 Studies contributing to the COVID-19 Host Genetics Initiative |
| Supplementary Table 3 Summary of Mendelian randomization methods and sensitivity analyses used |
| Supplementary Figure 1 Directed acyclic graph outlining Mendelian randomisation assumptions |
| Supplementary Figure 2 Flow chart of SNPs included in the main analysis of body mass index (BMI) on SARS-CoV-2/COVID-19 |
| Supplementary Figure 3 (a) Main analysis: Effect of body mass index (BMI) on SARS-CoV-2/ COVID-19 (b) No-relevance analysis': Effect of SARS-CoV-2/COVID-19 on BMI |
| Supplementary Figure 4 Effect of body mass index (BMI) on SARS-CoV-2/COVID-19: Inverse variance weighting (IVW) estimates from most recent GIANT BMI GWAS that includes UK Biobank (UKB) and the previous GWAS that excludes UKB |
| Supplementary Table 4 Cochran's Q test results of between SNP heterogeneity for the main the Mendelian randomisation (MR) association of effect of body mass index (BMI) on COVID-19 (4a), and zero-relevance test of potential 'biased' effect of COVID-19 and BMI (4b) |
| Supplementary Table 5 MR Egger intercept results for the MR associations between body mass index (BMI) and COVID-19, and between COVID-19 and BMI |
| Supplementary Figure 5 Population stratification: (a) Effect of body mass index (BMI) on skin tanning; (b) Effect of COVID-19 on skin tanning |
| Supplementary Table 6 SNPs associated with COVID-19 |
| Supplementary Figure 6. Forest plots of single SNP analysis of the association between COVID-19 and body mass index (BMI): for comparisons A1, A2 and B2 respectively (where there was some evidence of SNP heterogeneity) |

**Supplementary Table 1 Summary of one/two sample Mendelian randomisation (MR) studies that assessed the effect of body mass index (BMI) on SARS-CoV-2/COVID-19**

| **Author, year (PMID)** | **Type of MR** | **BMI GWAS** | **SARS-CoV-2/COVID-19 comparison groups** | **Sample sizes/ Release** | **Statistical analyses** | **Main results** | **Attempt to explore selection bias/ misclassification** |
|---|---|---|---|---|---|---|---|
| Gao et al, 2022 (35031696) | 2SMR | Yengo et al. | COVID-19 vs population (C2)<br><br>Hospitalised COVID-19 vs population (B2)<br><br>Very severe respiratory COVID-19 vs population (A2) | 38,984 COVID-19 cases/ 1,644,784 controls<br><br>9,986 hospitalised COVID-19 cases/ 1,877,672 controls<br><br>5,101 very severe respiratory COVID-19 cases / 1,383,241 controls<br><br>Release 5 (18th January 2021) | Main: IVW<br>Sensitivity: MR-Egger, weighted median, weighted mode<br><br>Observational: UKB | OR 1.20 (95% CI 1.13, 1.28) per 1 SD increase in BMI (C2)<br><br>OR 1.50 (95% CI 1.32, 1.70) per 1 SD increase in BMI (B2)<br><br>OR 1.44 (95% CI 1.19, 1.73) per 1 SD increase in BMI (A2) | No |
| Leong et al, 2021 (33661905) | 2SMR | Locke et al. | COVID-19 vs population (C2) – main analyis<br><br>Hospitalised COVID-19 vs population (B2) – main analysis<br><br>Very severe COVID-19 vs not hospitalised COVID-19 (A1)<br><br>Very severe COVID-19 vs poulation (A2) | 14,134 COVID-19 cases/ 1,284,876 controls<br><br>6,406 hospitalised COVID-19 cases/ 902,088 controls<br><br>269 very severe COVID-19 cases/ 688 controls<br><br>4,933 very severe COVID-19 cases/ 1,398,672 controls | Main: IVW<br>Sensitivity: weighted median, MR-Egger, mode-base estimate, MR-PRESSO, leave-one-out analysis | OR 1.29 (95% CI 1.10, 1.53) per 1 SD increase in BMI (C2)<br><br>OR: 1.69 (95% CI 1.31, 2.17) per 1 SD increase in BMI (B2)<br><br>OR 1.04 (95%CI 0.24, 4.50) per 1 SD increase in BMI (A1)<br><br>OR 1.67 (95%CI 1.24, 2.26) per 1 SD increase in BMI (A2) | Comparison of different case/control groups but not looking specifically at selection/ misclassification |

| | | | | | | | |
|---|---|---|---|---|---|---|---|
| | | | Hospitalised COVID-19 vs not hospitalised COVID-19 (B1)<br><br>COVID-19 vs lab/self-reported COVID-19 negative (C1)<br><br>Predicted COVID-19 vs predicted or self-reported COVID-19 negative (D1) | 2,430 hospitalised COVID-19 cases/ 8,488 controls<br><br>24,057 COVID-19 cases / 218,062 controls<br><br>3,204 COVID-19 cases/ 35,728 controls<br><br>Release 4 (20th October 2020) | | OR 1.50 (95% CI 0.95, 2.37) per 1 SD increase in BMI (B1)<br><br>OR 1.21 (95% CI 1.02, 1.44) per 1 SD increase in BMI (C1)<br><br>OR (0.93, 95%CI 0.71, 1.21) per 1 SD increase in BMI (D1) | |
| Li S et al, 2021 (33536004) | 2SMR | Yengo et al. | Very severe respiratory COVID-19 vs population (A2)<br><br>Hospitalised COVID-19 vs population (B2) | 2,972 cases / 284,472 controls<br><br>6,492 cases / 1,012,809 controls<br><br>Release 4 (September 2020) | Main: IVW<br>Sensitivity: MR-Egger, weighted median, weighted mode | OR 1.91 (95% CI 1.55, 2.35) per 1 SD increase in BMI (A2)<br><br>OR 1.75 (95% CI 1.52, 2.01) per 1 SD increase in BMI (B2) | No |
| Lorincz-Comi et al, 2021 (33846372) | 2SMR | Yengo et al. | COVID-19 hospitalised vs not hospitalised COVID-19 (B1) | 928 cases / 2,028 controls (EUR)<br><br>2,430 cases/ 8,378 controls (Mixed)<br><br>Release 3 for EUR (2nd July 2020) and 4 for mixed (20th October 2020) | Main: IVW<br>Sensitivity: MRMix, IMRP, MR-Egger | OR 0.89 (95% CI 0.62, 1.28) per 1 SD increase in BMI (EUR)<br><br>OR 1.12 (95% CI 0.90, 1.38) per 1 SD increase in BMI (Mixed) | No |
| Freuer et al, 2021 (33631142) | 2SMR | Pulit et al. | COVID-19 vs population (C2) | 6,696 cases/ 1,073,072 controls) | Main: radial IVW, random-effects IVW<br>Sensitivity: MR-PRESSO, RAPS, | OR 1.31 (95%CI 1.15, 1.50) per 1 SD increase in BMI (C2) | No |

| | | | | | | | |
|---|---|---|---|---|---|---|---|
| | | | Hospitalised COVID-19 vs population (B2) | 3,199 cases/ 897,488 controls<br><br>Release 3 (29th June 2020) | weighted mean, weigted mode, MR-Egger | OR 1.62 (95% CI 1.32, 1.99) per 1 SD increase in BMI (B2) | |
| Ponsford et al, 2020 (32966752) | 2SMR | Yengo et al. | Severe COVID-19 vs controls (with no or mild COVID-19 symptoms)<br><br>Hospitalised COVID-19 vs population (B2) | 1,610 cases/ 2,205 controls in Italy and Spain<br><br>3,199 cases/ 897,488 controls<br><br>Release 3 (29th June 2020) | Main: IVW<br>Sensitivity: MR-Egger, weighted median | OR: 1.75 (95% CI 1.20, 2.57) per 1 SD increase in BMI (severe COVID-19)<br><br>OR 1.47 (95% CI 1.18, 1.83) per 1 SD increase in BMI (B2) | |
| Aung et al, 2020 (33262790) | 1SMR and 2SMR | Locke et al. | COVID-19 vs controls (tested negative or untested) in UK Biobank<br><br>COVID-19 vs population (C2) | 1,211 cases / 387,079 controls<br>16th March 2020 – 31st May 2020<br><br>1,678 cases/ 674,635 controls<br><br>Release 2 (15th May 2020) | Main: IVW<br>Sensitivity: MR-Egger, weighted median, MR-PRESSO | OR 1.15 (1.05, 1.36) per 1 SD increase in BMI (UKB)<br><br>OR 1.80 (95% CI 1.28, 2.54) per 1 SD increase in BMI (C2) | Sensitivity analysis in 1SMR restricted to only those tested for SARS-CoV-2 |

EUR: Europeans; IMRP: Iterative Mendelian Randomization and Pleiotropy approach; IVW: inverse-variance weighting, MRMix: Mendelian randomisation using mixture models; MR-PRESSO: Mendelian Randomisation Pleiotropy RESidual Sum and Outlier

**Supplementary Table 2 Studies contributing to the COVID-19 Host Genetics Initiative**

| Group/studies | Cases | Controls |
|---|---|---|
| **A1: Very severe respiratory confirmed COVID-19 vs not hospitalised COVID-19** | | |
| BQC19_EUR | 55 | 151 |
| BoSCO_EUR | 59 | 262 |
| FinnGen_FIN | 54 | 224 |
| SPGRX_EUR | 101 | 51 |
| **A2: Very severe respiratory confirmed COVID-19 vs population** | | |
| ANCESTRY_Freeze_Four_EUR | 667 | 113882 |
| Amsterdam_UMC_COVID_study_group_EUR | 66 | 1413 |
| BQC19_EUR | 165 | 743 |
| BRACOVID_AMR | 539 | 1149 |
| BelCovid_EUR | 184 | 1477 |
| CHULACOVID_EAS | 137 | 326 |
| CU_AFR | 133 | 2610 |
| CU_EUR | 203 | 2149 |
| Egypt_hgCOVID_hub_AFR | 167 | 180 |
| FinnGen_FIN | 190 | 278193 |
| GENCOVID_EUR | 1077 | 2443 |
| GHS_Freeze_145_EUR | 321 | 108168 |
| GNH_SAS | 151 | 34013 |
| HOSTAGE_EUR | 1779 | 9116 |
| INMUNGEN_CoV2_EUR | 70 | 654 |
| JapanTaskForce_EAS | 155 | 1705 |
| PMBB_AFR | 55 | 8622 |
| SPGRX_EUR | 101 | 302 |
| SaudiHumanGenomeProgram_ARAB | 147 | 768 |
| SweCovid_EUR | 113 | 3748 |
| UKBB_EUR | 417 | 420114 |
| Vanda_EUR | 58 | 900 |
| genomicc_EAS | 149 | 745 |
| genomicc_EUR | 1676 | 8380 |
| idipaz24genetics_EUR | 59 | 75 |
| **B1: Hospitalised COVID-19 vs not hospitalised COVID-19** | | |
| ANCESTRY_Freeze_Four_AFR | 135 | 1494 |
| ANCESTRY_Freeze_Four_AMR | 199 | 3553 |
| ANCESTRY_Freeze_Four_EUR | 1484 | 23869 |
| BelCovid_EUR | 365 | 121 |
| BoSCO_EUR | 212 | 512 |
| CHOP_CAG_AFR | 147 | 412 |
| DECODE_EUR | 89 | 1808 |
| EstBB_EUR | 271 | 2335 |
| FHoGID_EUR | 470 | 280 |
| GENCOVID_EUR | 1267 | 331 |
| GHS_Freeze_145_EUR | 773 | 4503 |
| GNH_SAS | 339 | 4812 |

| | | |
|---|---|---|
| Generation_Scotland_EUR | 554 | 3674 |
| IranCovid_MID | 210 | 183 |
| MVP_AFR | 1300 | 3573 |
| MVP_AMR | 517 | 1962 |
| MVP_EUR | 2417 | 9251 |
| PMBB_AFR | 66 | 100 |
| QGP_ARAB | 60 | 640 |
| SPGRX_EUR | 311 | 51 |
| UKBB_AFR | 101 | 291 |
| UKBB_CSA | 126 | 489 |
| UKBB_EUR | 3067 | 8947 |
| **B2: Hospitalised COVID-19 vs population** | | |
| ANCESTRY_Freeze_Four_EUR | 1484 | 113882 |
| Amsterdam_UMC_COVID_study_group_EUR | 108 | 1413 |
| BQC19_EUR | 378 | 530 |
| BRACOVID_AMR | 853 | 835 |
| BelCovid_EUR | 367 | 1477 |
| CHULACOVID_EAS | 137 | 326 |
| CU_AFR | 304 | 2610 |
| CU_EUR | 453 | 2149 |
| Egypt_hgCOVID_hub_AFR | 167 | 180 |
| FinnGen_FIN | 273 | 278193 |
| GENCOVID_EUR | 1287 | 2443 |
| GHS_Freeze_145_EUR | 773 | 108168 |
| GNH_SAS | 339 | 33825 |
| HOSTAGE_EUR | 3151 | 12483 |
| JapanTaskForce_EAS | 572 | 1705 |
| PMBB_AFR | 126 | 8551 |
| SPGRX_EUR | 311 | 302 |
| SaudiHumanGenomeProgram_ARAB | 147 | 768 |
| SweCovid_EUR | 113 | 3748 |
| UKBB_EUR | 3067 | 417464 |
| Vanda_EUR | 58 | 900 |
| genomicc_EAS | 149 | 745 |
| genomicc_EUR | 1676 | 8380 |
| idipaz24genetics_EUR | 106 | 75 |
| **C1: COVID-19 vs lab/self-reported negative** | | |
| 23ANDME_AFR | 506 | 3110 |
| 23ANDME_EUR | 9913 | 85072 |
| 23ANDME_HIS | 2553 | 13086 |
| ANCESTRY_Freeze_Four_EUR | 2417 | 14933 |
| BQC19_EUR | 206 | 327 |
| DECODE_EUR | 1897 | 29014 |
| EstBB_EUR | 313 | 12019 |
| GNH_SAS | 114 | 256 |
| INTERVAL_EUR | 161 | 1119 |
| Lifelines_EUR | 358 | 1253 |
| MGI_EUR | 122 | 508 |

| | | |
|---|---|---|
| MVP_AFR | 1217 | 9204 |
| MVP_AMR | 510 | 3325 |
| MVP_EUR | 1520 | 24135 |
| NTR_EUR | 145 | 117 |
| PHBB_AFR | 60 | 375 |
| PHBB_EUR | 151 | 3118 |
| PHBB_HIS | 66 | 276 |
| PMBB_AFR | 166 | 934 |
| UKBB_AFR | 68 | 268 |
| UKBB_CSA | 66 | 350 |
| UKBB_EUR | 1310 | 13646 |
| genomicsengland100kgp_EUR | 218 | 1617 |
| **C2: COVID-19 vs population** | | |
| ACCOuNT_AFR | 57 | 799 |
| ANCESTRY_Freeze_Four_AFR | 1629 | 5688 |
| ANCESTRY_Freeze_Four_AMR | 3752 | 12323 |
| ANCESTRY_Freeze_Four_EAS | 120 | 570 |
| ANCESTRY_Freeze_Four_EUR | 25353 | 113882 |
| Amsterdam_UMC_COVID_study_group_EUR | 108 | 1413 |
| BQC19_EUR | 477 | 431 |
| BRACOVID_AMR | 853 | 835 |
| BelCovid_EUR | 488 | 1477 |
| BioVU_EUR | 141 | 70615 |
| CCHC_AMR_HIS | 95 | 3140 |
| CCPM_AMR | 220 | 3111 |
| CCPM_EUR | 896 | 24851 |
| CHULACOVID_EAS | 137 | 326 |
| CU_AFR | 332 | 2610 |
| CU_EUR | 508 | 2149 |
| Corea_EAS | 108 | 6500 |
| Coronagenes_EUR | 181 | 734 |
| DECODE_EUR | 4256 | 270934 |
| EXCEED_EUR | 236 | 1083 |
| Egypt_hgCOVID_hub_AFR | 167 | 180 |
| EraCORE_EUR | 302 | 1646 |
| EstBB_EUR | 4070 | 189406 |
| FinnGen_FIN | 2695 | 278193 |
| GCAT_EUR | 447 | 4541 |
| GENCOVID_EUR | 1697 | 2443 |
| GFG_EUR | 136 | 4978 |
| GHS_Freeze_145_AFR | 128 | 3050 |
| GHS_Freeze_145_AMR | 88 | 1307 |
| GHS_Freeze_145_EUR | 5276 | 108168 |
| GNH_SAS | 5151 | 29013 |
| Generation_Scotland_EUR | 1712 | 17525 |
| Genetics_COVID19_Korea_EAS | 624 | 6549 |
| Genotek_EUR | 676 | 12317 |
| HOSTAGE_EUR | 3151 | 12483 |

| | | |
|---|---|---|
| HUNT_EUR | 283 | 52072 |
| Helix_EUR | 178 | 5441 |
| INMUNGEN_CoV2_EUR | 236 | 654 |
| INTERVAL_EUR | 2098 | 39733 |
| JapanTaskForce_EAS | 614 | 1705 |
| LGDB_EUR | 275 | 1313 |
| Lifelines_EUR | 895 | 26600 |
| MGI_AFR | 111 | 2989 |
| MGI_EUR | 122 | 51458 |
| MOBA_EUR | 391 | 56038 |
| MVP_AFR | 4893 | 94556 |
| MVP_AMR | 2497 | 41100 |
| MVP_EUR | 11778 | 357198 |
| NTR_EUR | 228 | 5265 |
| PMBB_AFR | 379 | 8298 |
| PMBB_EUR | 60 | 9702 |
| QGP_ARAB | 700 | 13360 |
| SINAI_COVID_EUR | 942 | 3784 |
| SPGRX_EUR | 362 | 302 |
| SaudiHumanGenomeProgram_ARAB | 147 | 768 |
| Stanford_EUR | 169 | 190 |
| SweCovid_EUR | 113 | 3748 |
| TOPMed_CHRIS10K_EUR | 92 | 2373 |
| TOPMed_Gardena_EUR | 452 | 458 |
| TwinsUK_EUR | 247 | 2320 |
| UKBB_AFR | 426 | 6210 |
| UKBB_AMR | 51 | 929 |
| UKBB_CSA | 695 | 8181 |
| UKBB_EAS | 104 | 2605 |
| UKBB_EUR | 13256 | 407275 |
| UKBB_MID | 89 | 1510 |
| Vanda_EUR | 58 | 900 |
| WGHS_EUR | 225 | 11887 |
| genomicc_EAS | 149 | 745 |
| genomicc_EUR | 1676 | 8380 |
| genomicsengland100kgp_EUR | 822 | 44369 |
| genomicsengland100kgp_SAS | 235 | 4205 |
| idipaz24genetics_EUR | 106 | 75 |
| thaicovid_EAS | 191 | 113 |
| **D1: predicted COVID-19 from self-reported symptoms vs predicted or self-reported non COVID-19** | | |
| GS_EUR | 132 | 3610 |
| GeneRISK_EUR | 154 | 3451 |
| Helix_EUR | 605 | 4778 |
| Lifelines_EUR | 1427 | 16833 |
| NTR_EUR | 603 | 4669 |
| RS_EUR | 283 | 2387 |

**Supplementary Table 3 Summary of Mendelian randomization methods and sensitivity analyses used**

<table>
<tr><th>Method</th><th colspan="2">Description</th></tr>
<tr><td colspan="3">Main analyses</td></tr>
<tr><td>Mendelian randomisation (MR)</td><td colspan="2">An instrumental variable approach that uses genetic variants as instruments for a modifiable exposure. MR is less prone to bias by confounding and reverse causality due to the random allocation of genetic variants at conception. However, a number of strong, often unverifiable assumptions are required. Genetic instruments must 1) be robustly and statistically strongly associated with the risk factor (Relevance assumption), 2) be independent of confounders between it and the outcome (Independence assumption) and 3) only be associated with the outcome via their effect on the risk factor (Exclusion restriction assumption).</td></tr>
<tr><td>Inverse variance weighted (IVW)</td><td colspan="2">Estimates a causal effect of the exposure on the outcome by regressing the SNP-exposure association on the SNP-outcome association and constraining the intercept of the regression to zero. Assumes that in the presence of horizontal pleiotropic paths there is no correlation between the association of the SNP-exposure (BMI) and SNP-pleiotropic path (also known as the Instrument Strength Independent of Direct Effect (InSIDE) assumption).</td></tr>
<tr><td colspan="3">Methods to explore MR assumptions</td></tr>
<tr><td>F-statistic</td><td>Relevance assumption and weak instrument bias</td><td>In the two-sample summary data context and with uncorrelated genetic variants, the F-statistic for variant j can be approximated as $F_j = beta^2 / se^2$. Higher F-statistic values suggest lower risk of weak instrument bias.</td></tr>
<tr><td>MR-Egger</td><td>Horizontal pleiotropy</td><td>Identical to IVW, except that it does not constrain the regression line to go through zero. This means that the slope of the MR-Egger regression represents the estimate of the causal effect controlling for unbalanced horizontal pleiotropy. A non-null MR-Egger intercept provides evidence of horizontal pleiotropy. However, MR-Egger has less statistical power than IVW and there should be no violation of the InSIDE assumption</td></tr>
<tr><td>Cochran's Q statistics</td><td>Horizontal pleiotropy</td><td>Tests for between-SNP heterogeneity</td></tr>
<tr><td>Weighted median (WM)</td><td>Horizontal pleiotropy</td><td>Provides an unbiased estimate when up to 50% of the SNPs used in the instrument violate the IV assumptions</td></tr>
<tr><td>Sample definition/ underlying population</td><td>Confounding (via population stratification)</td><td>Confounding of SNP-outcome associations (Independence assumption) can occur where there is population substruction within the study. To minimise this bias, analyses are carried out using</td></tr>
</table>

| | | |
|---|---|---|
| | | GWASs from the same underlying population (e.g., both from European ancestry). |
| Independent GWAS samples | Sample overlap | If samples overlap, then features of two-sample MR, such as weak instrument bias being expected to be towards the null, become more like one-sample MR, particularly with substantial overlap. |

**Supplementary Figure 1 Directed acyclic graph outlining Mendelian randomisation assumptions**

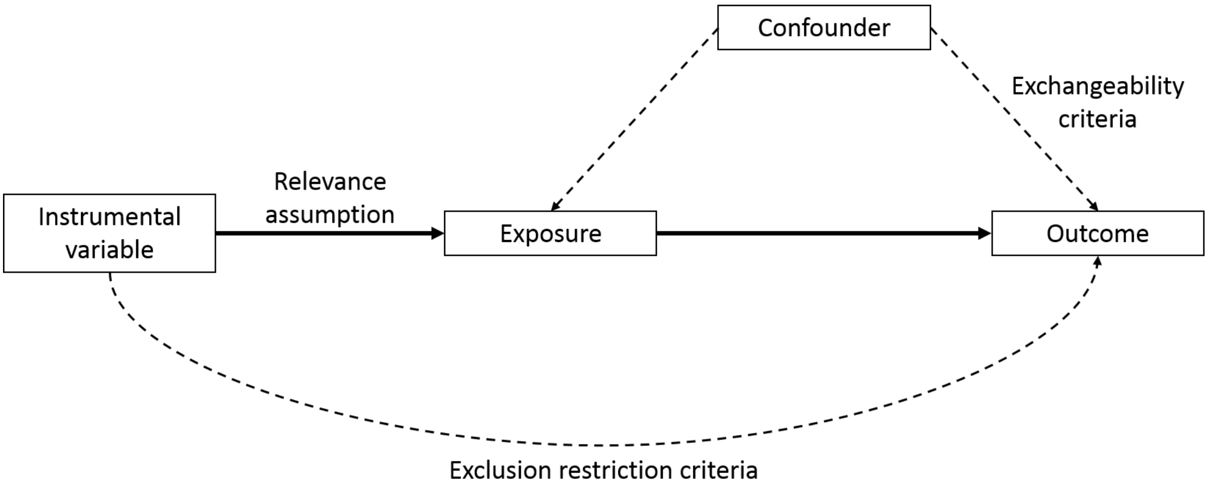

**Supplementary Figure 2  Flow chart of SNPs included in the main analysis of body mass index (BMI) on SARS-CoV-2/COVID-19**

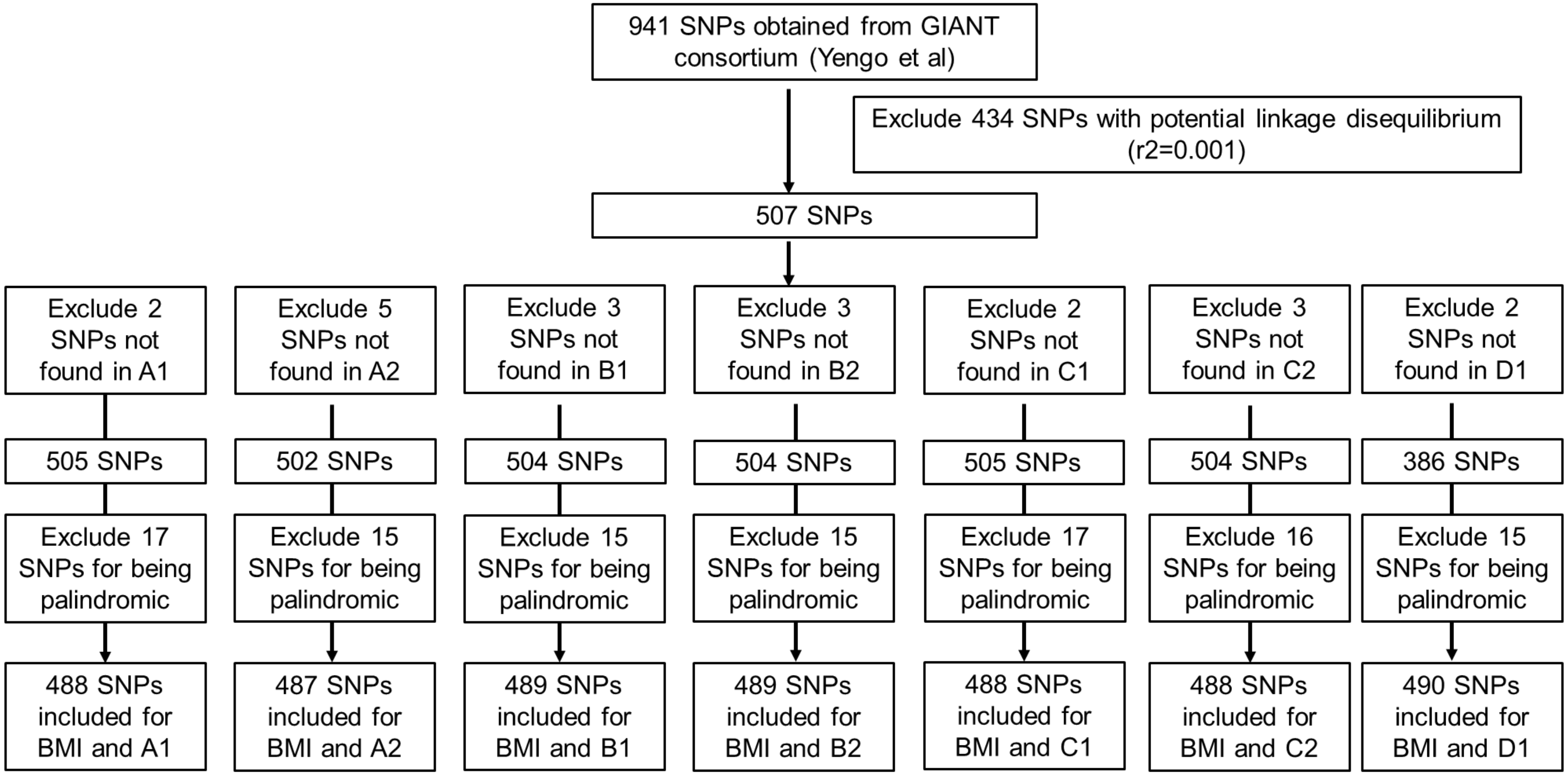


SNPs= single-nucleotide polymorphisms; GIANT=Genetic Investigation of ANthropometric Traits

A1: very severe respiratory confirmed COVID-19 vs not hospitalised COVID-19; A2: very severe respiratory confirmed COVID-19 vs population; B1: hospitalised COVID-19 vs not hospitalised COVID-19; B2: hospitalised COVID-19 vs population; C1: COVID-19 vs lab/self-reported negative; C2: COVID-19 vs population; D1: predicted COVID-19 from self-reported symptoms vs predicted or self-reported non COVID-19.
For comparisons A1, C1 and D1, one proxy SNP was included.

**Supplementary Figure 3 (a) Main analysis: Effect of body mass index (BMI) on SARS-CoV-2/ COVID-19 (b) No-relevance analysis': Effect of SARS-CoV-2/COVID-19 on BMI**

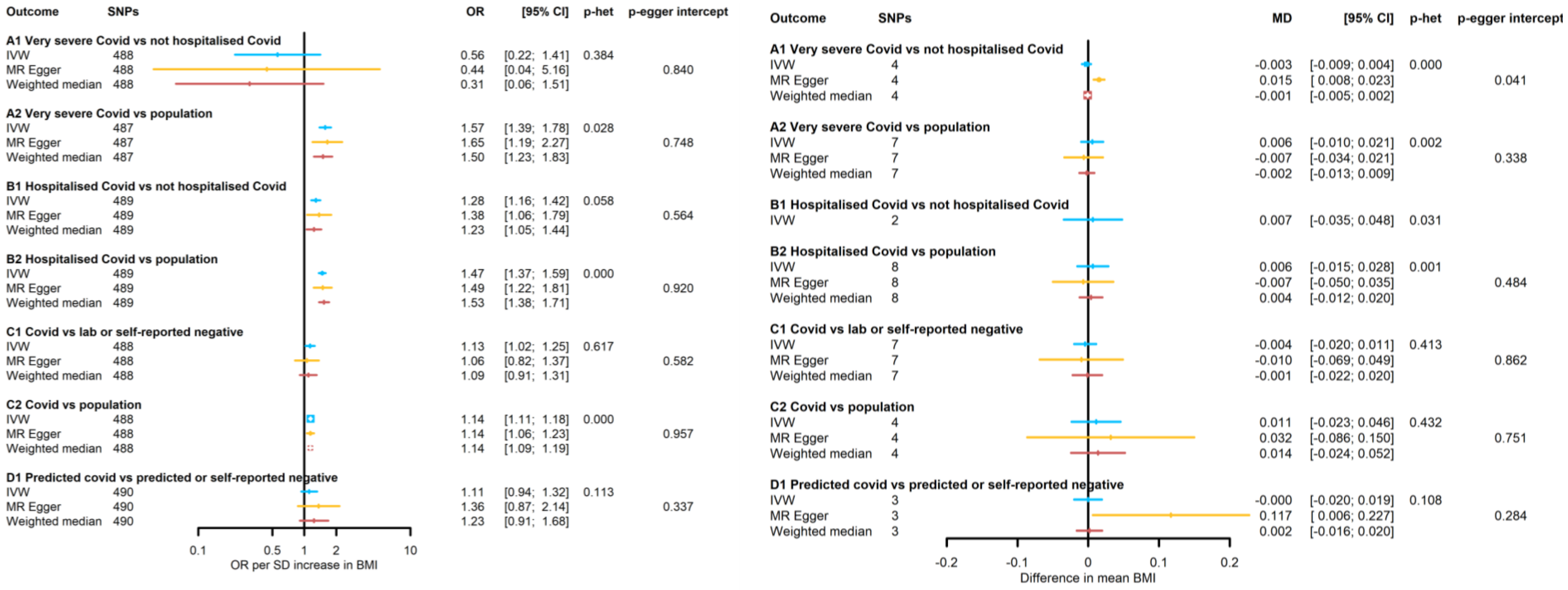


Footnote: IVW: inverse variance weighting; SNPs: single nuclear polymorphism. Groups A1, C1 and D1 are from version 4 whilst the others are from version 6. Note that different studies are contributing to each case/control group (see **Supplementary Table 2**), p-het: p-value from Cochranes Q statistic assessing SNP heterogeneity (see **Supplementary Table 4** for Cochranes Q statistic); p-egger intercept: p-value for MR Egger intercept (see **Supplementary Table 5** for intercept and standard error). For the 'no relevance' analysis, for groups A1, C1 and D1 a threshold of $p<5x10^{-6}$ was used for selection of SNPs; C1 includes one proxy SNP, and D1 includes 2 proxy SNPs.

**Supplementary Figure 4 Effect of body mass index (BMI) on SARS-CoV-2/COVID-19: Inverse variance weighting (IVW) estimates from most recent GIANT BMI GWAS that includes UK Biobank (UKB) and the previous GWAS that excludes UKB**

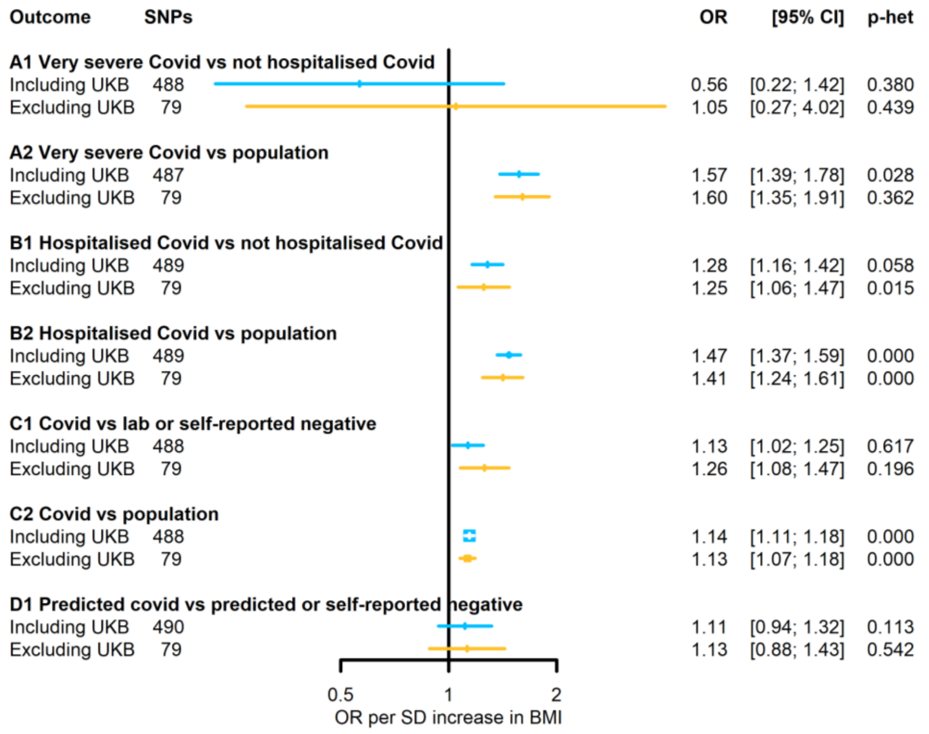


SNPs: single nuclear polymorphism, p-het: p-value from Cochranes Q statistic assessing SNP heterogeneity

**Supplementary Table 4 Cochran's Q test results of between SNP heterogeneity for the main the Mendelian randomisation (MR) association of effect of body mass index (BMI) on COVID-19 (4a), and zero-relevance test of potential 'biased' effect of COVID-19 and BMI (4b)**

| Covid group | Method | Q test | df | p-value |
|---|---|---|---|---|
| **4a Main analyses (BMI on COVID-19)** | | | | |
| A1: Very severe COVID-19 vs not hospitalised COVID-19 | IVW | 495.6 | 487 | 0.384 |
| | MR Egger | 495.5 | 486 | 0.373 |
| A2: Very severe COVID-19 vs population | IVW | 547.2 | 486 | 0.028 |
| | MR Egger | 547.1 | 485 | 0.026 |
| B1: Hospitalised COVID-19 vs not hospitalised COVID-19 | IVW | 538.1 | 488 | 0.058 |
| | MR Egger | 537.7 | 487 | 0.055 |
| B2: Hospitalised COVID-19 vs population | IVW | 608.7 | 488 | 1.56E-04 |
| | MR Egger | 608.7 | 487 | 1.38E-04 |
| C1: COVID-19 vs lab or self-reported negative | IVW | 477.1 | 487 | 0.617 |
| | MR Egger | 476.8 | 486 | 0.609 |
| C2: COVID-19 vs population | IVW | 618.8 | 487 | 4.56E-05 |
| | MR Egger | 618.8 | 486 | 4.02E-05 |
| D1: Predicted COVID-19 vs predicted or self-reported | IVW | 527.1 | 489 | 0.113 |
| negative | MR Egger | 526.1 | 488 | 0.113 |
| **4b 'No-relevance analyses' (MR of COVID-19 on BMI)** | | | | |
| A1: Very severe COVID-19 vs not hospitalised COVID-19 | IVW | 23.2 | 3 | <0.001 |
| | MR Egger | 0.2 | 2 | 0.890 |
| A2: Very severe COVID-19 vs population | IVW | 21.4 | 6 | 0.002 |
| | MR Egger | 17.5 | 5 | 0.004 |
| B1: Hospitalised COVID-19 vs not hospitalised COVID-19 | IVW | 4.7 | 1 | 0.031 |
| B2: Hospitalised COVID-19 vs population | IVW | 23.9 | 7 | 0.001 |
| | MR Egger | 21.9 | 6 | 0.001 |
| C1: COVID-19 vs lab or self-reported negative | IVW | 6.1 | 6 | 0.413 |
| | MR Egger | 6.1 | 5 | 0.301 |
| C2: COVID-19 vs population | IVW | 2.8 | 3 | 0.432 |
| | MR Egger | 2.6 | 2 | 0.275 |
| D1: Predicted COVID-19 vs predicted or self-reported | IVW | | | |
| negative | | 4.5 | 2 | 0.108 |
| | MR Egger | 0.1 | 1 | 0.790 |

**Supplementary Table 5 MR Egger intercept results for the MR associations between body mass index (BMI) and COVID-19, and between COVID-19 and BMI**

| Description | MR Egger intercept | SE | p-value |
|---|---|---|---|
| **Main analysis (MR of BMI on COVID-19)** | | | |
| A1: Very severe COVID-19 vs not hospitalised COVID-19 | 0.004 | 0.020 | 0.840 |
| A2: Very severe COVID-19 vs population | -0.001 | 0.003 | 0.748 |
| B1: Hospitalised COVID-19 vs not hospitalised COVID-19 | -0.001 | 0.002 | 0.564 |
| B2: Hospitalised COVID-19 vs population | 0.000 | 0.002 | 0.920 |
| C1: COVID-19 vs lab or self-reported negative | 0.001 | 0.002 | 0.582 |
| C2: COVID-19 vs population | 0.000 | 0.001 | 0.957 |
| D1: Predicted COVID-19 vs predicted or self-reported negative | -0.004 | 0.004 | 0.337 |
| **'No-relevance analysis' (MR of COVID-19 on BMI)** | | | |
| A1: Very severe COVID-19 vs not hospitalised COVID-19 | -0.022 | 0.005 | 0.041 |
| A2: Very severe COVID-19 vs population | 0.003 | 0.003 | 0.338 |
| B2: Hospitalised COVID-19 vs population | 0.002 | 0.003 | 0.484 |
| C1: COVID-19 vs lab or self-reported negative | -0.002 | 0.003 | 0.575 |
| C2: COVID-19 vs population | -0.001 | 0.003 | 0.751 |
| D1: Predicted COVID-19 vs predicted or self-reported negative | -0.023 | 0.011 | 0.284 |

Note: it was not possible to estimate MR Egger intercept for group B1 in the 'no-relevance' analysis due to the small number of instruments (less than 3)

**Supplementary Figure 5 Population stratification: (a) Effect of body mass index (BMI) on skin tanning; (b) Effect of COVID-19 on skin tanning**

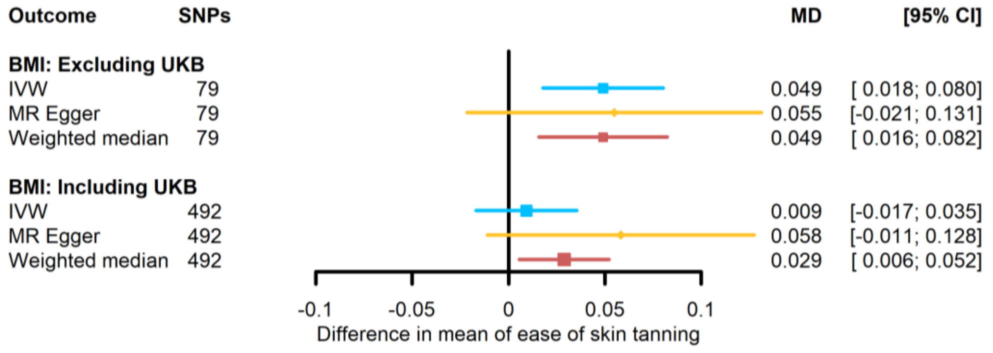


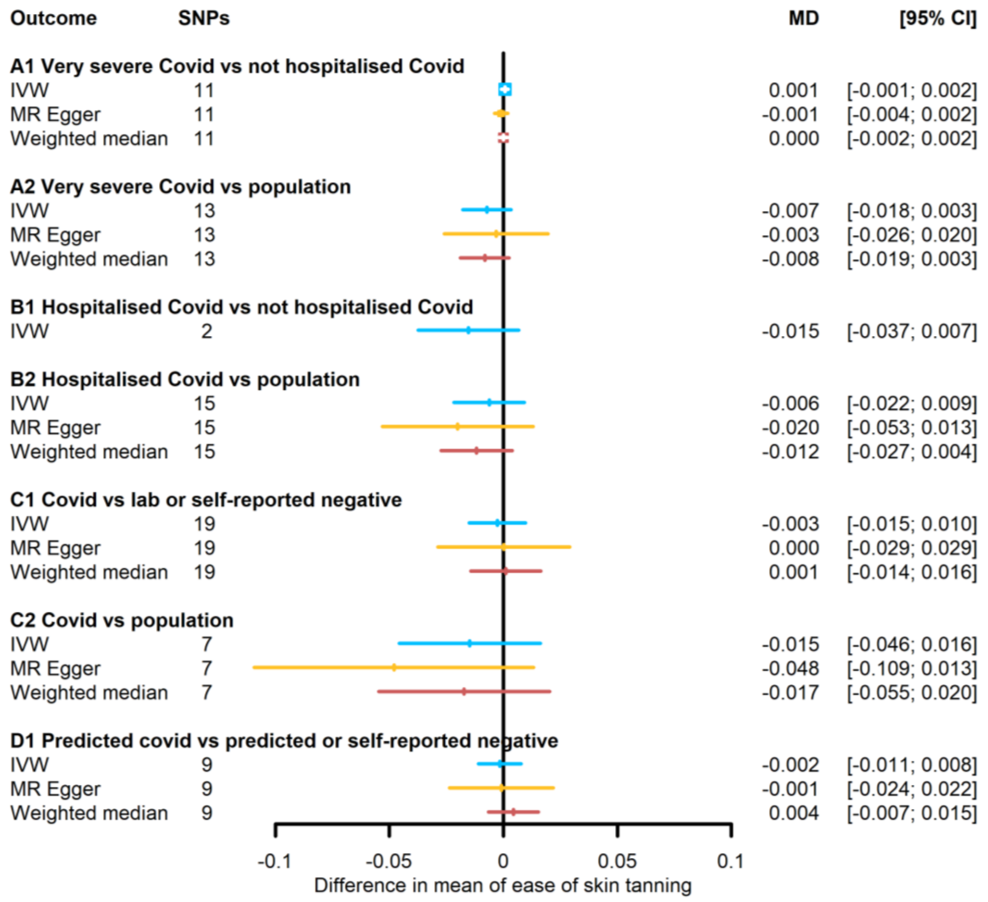


IVW: inverse variance weighting; SNPs: single nuclear polymorphism, MD: Mean difference (also referred to as Difference in mean); CI: Confidence interval

**Supplementary Table 6 SNPs associated with COVID-19**

| Covid group | Sample size | rsID | effect allele | other allele | EAF | logOR | SE | P-value | Pseudo F-statistics | $R^2$ |
|---|---|---|---|---|---|---|---|---|---|---|
| A1: Very severe Covid vs not hospitalised Covid | 957 | rs145285136 | A | G | 0.016 | 4.147 | 0.853 | 1.163E-06 | 23.6 | 0.024 |
| | 957 | rs12069768 | T | C | 0.040 | 1.838 | 0.394 | 3.101E-06 | 21.8 | 0.022 |
| | 957 | rs4462982 | C | T | 0.542 | 0.731 | 0.154 | 2.081E-06 | 22.5 | 0.023 |
| | 957 | rs545631184 | T | C | 0.072 | 2.362 | 0.508 | 3.304E-06 | 21.6 | 0.022 |
| | 957 | rs73064425 | T | C | 0.110 | 1.406 | 0.297 | 2.213E-06 | 22.4 | 0.023 |
| | 957 | rs1107783 | C | T | 0.261 | -0.889 | 0.193 | 4.284E-06 | 21.1 | 0.022 |
| | 957 | rs12206024 | A | G | 0.188 | 0.896 | 0.194 | 3.916E-06 | 21.3 | 0.022 |
| | 957 | rs59492037 | G | A | 0.100 | 1.283 | 0.259 | 7.072E-07 | 24.6 | 0.025 |
| | 957 | rs112338868 | C | T | 0.016 | 4.242 | 0.838 | 4.114E-07 | 25.6 | 0.026 |
| | 957 | rs1522633 | A | T | 0.375 | 0.809 | 0.157 | 2.76E-07 | 26.4 | 0.027 |
| | 957 | rs3812875 | G | A | 0.037 | 1.771 | 0.381 | 3.437E-06 | 21.6 | 0.022 |
| Harmonised SNPs | | | | | | | | | 23.8 | 0.1059 |
| When assuming a prevalence of 8%, total $R^2$=0.14 | | | | | | | | | | |
| A2: Very severe Covid vs population | 1,010,654 | rs10774671 | A | G | 0.668 | 0.126 | 0.019 | 3.60E-11 | 43.8 | 4.34E-05 |
| | 1,010,191 | rs111837807 | C | T | 0.09793 | 0.204 | 0.03 | 2.15E-11 | 44.8 | 4.44E-05 |
| | 996,978 | rs12259804 | C | T | 0.07149 | -0.275 | 0.05 | 4.87E-08 | 29.8 | 2.99E-05 |
| | 992,464 | rs12660421 | A | G | 0.03706 | 0.355 | 0.058 | 1.14E-09 | 37.1 | 3.73E-05 |
| | 1,009,233 | rs13050728 | C | T | 0.6563 | -0.167 | 0.02 | 1.24E-16 | 68.5 | 6.79E-05 |
| | 1,010,654 | rs1405655 | C | T | 0.3229 | 0.114 | 0.019 | 4.06E-09 | 34.6 | 3.42E-05 |
| | 975,766 | rs2277732 | A | C | 0.3153 | 0.203 | 0.021 | 8.48E-23 | 96.6 | 9.90E-05 |
| | 959,347 | rs34536443 | C | G | 0.03966 | 0.394 | 0.06 | 4.89E-11 | 43.2 | 4.51E-05 |
| | 1,009,297 | rs35154152 | C | T | 0.08958 | -0.205 | 0.031 | 2.36E-11 | 44.7 | 4.42E-05 |
| | 1,009,760 | rs35731912 | T | C | 0.08046 | 0.588 | 0.031 | 4.25E-81 | 363.8 | 3.60E-04 |
| | 1,010,251 | rs495828 | G | T | 0.7857 | -0.134 | 0.022 | 6.56E-10 | 38.1 | 3.78E-05 |
| | 996,635 | rs71479671 | A | G | 0.4598 | -0.131 | 0.024 | 4.19E-08 | 30.1 | 3.02E-05 |
| | 1,007,437 | rs8080583 | A | C | 0.1789 | -0.127 | 0.023 | 2.08E-08 | 31.4 | 3.12E-05 |

| | | | | | | | | | | |
|---|---|---|---|---|---|---|---|---|---|---|
| Harmonised SNPs | | | | | | | | | 88.0 | 0.0006 |
| When assuming a prevalence of 4%, total $R^2$=0.015 | | | | | | | | | | |
| B1: Hospitalised Covid vs not hospitalised Covid | 87,671 | rs11088247 | G | C | 0.3304 | 0.095 | 0.016 | 6.31E-09 | 33.7 | 3.85E-04 |
| | 87,505 | rs17713054 | A | G | 0.09075 | 0.304 | 0.028 | 2.04E-28 | 122.2 | 0.0014 |
| Harmonised SNPs | | | | | | | | | 78.0 | 0.0018 |
| When assuming a prevalence of 10%, total $R^2$=0.0028 | | | | | | | | | | |
| B2: Hospitalised Covid vs population | 2,085,803 | rs10774679 | T | C | 0.6505 | 0.076 | 0.011 | 4.70E-12 | 47.8 | 2.29E-05 |
| | 2,071,920 | rs111837807 | C | T | 0.08627 | 0.113 | 0.018 | 9.46E-10 | 37.4 | 1.81E-05 |
| | 2,078,167 | rs11702230 | T | C | 0.1174 | 0.1 | 0.017 | 3.22E-09 | 35.0 | 1.69E-05 |
| | 2,084,590 | rs12809318 | C | T | 0.52 | -0.063 | 0.011 | 9.53E-09 | 32.9 | 1.58E-05 |
| | 2,083,632 | rs13050728 | C | T | 0.6568 | -0.107 | 0.011 | 4.07E-21 | 88.9 | 4.27E-05 |
| | 2,085,803 | rs1405655 | C | T | 0.3324 | 0.075 | 0.011 | 1.12E-11 | 46.1 | 2.21E-05 |
| | 2,050,749 | rs2277732 | A | C | 0.2932 | 0.117 | 0.012 | 6.19E-22 | 92.7 | 4.52E-05 |
| | 2,019,040 | rs34536443 | C | G | 0.03888 | 0.174 | 0.031 | 1.82E-08 | 31.7 | 1.57E-05 |
| | 2,071,167 | rs35508621 | C | T | 0.07643 | 0.389 | 0.019 | 6.09E-89 | 399.8 | 1.93E-04 |
| | 1,790,208 | rs35705950 | T | G | 0.09959 | -0.112 | 0.02 | 1.22E-08 | 32.4 | 1.81E-05 |
| | 2,085,803 | rs505922 | T | C | 0.6534 | -0.1 | 0.011 | 2.09E-20 | 85.7 | 4.11E-05 |
| | 1,794,016 | rs61667602 | C | T | 0.1743 | -0.094 | 0.014 | 9.13E-12 | 46.5 | 2.59E-05 |
| | 2,084,446 | rs67579710 | A | G | 0.1057 | -0.103 | 0.017 | 1.03E-09 | 37.3 | 1.79E-05 |
| | 2,072,069 | rs766826 | T | C | 0.3239 | -0.082 | 0.012 | 1.15E-11 | 46.0 | 2.22E-05 |
| | 1,905,431 | rs9367106 | C | G | 0.03992 | 0.202 | 0.031 | 3.40E-11 | 43.9 | 2.31E-05 |
| Harmonised SNPs | | | | | | | | | 99.2 | 0.0004 |
| When assuming a prevalence of 3%, total $R^2$=0.0067 | | | | | | | | | | |
| C1: Covid vs lab or self-reported negative | 110624 | rs7535387 | G | C | 0.113 | 0.350 | 0.076 | 4.599E-06 | 21.0 | 0.000 |
| | 110624 | rs4656878 | A | G | 0.470 | 0.095 | 0.021 | 3.907E-06 | 21.3 | 0.000 |
| | 110624 | rs73062394 | T | A | 0.106 | 0.230 | 0.036 | 1.876E-10 | 40.6 | 0.000 |
| | 110624 | rs144636229 | G | A | 0.081 | 0.320 | 0.070 | 4.683E-06 | 21.0 | 0.000 |
| | 110624 | rs4974756 | C | G | 0.401 | -0.099 | 0.022 | 4.084E-06 | 21.2 | 0.000 |
| | 110624 | rs116335024 | T | C | 0.079 | 0.346 | 0.073 | 2.291E-06 | 22.3 | 0.000 |
| | 110624 | rs4130109 | A | T | 0.800 | -0.109 | 0.022 | 8.477E-07 | 24.2 | 0.000 |
| | 110624 | rs72691535 | A | C | 0.112 | 0.173 | 0.034 | 3.055E-07 | 26.2 | 0.000 |

| | | | | | | | | | | |
|---|---|---|---|---|---|---|---|---|---|---|
| | 110624 | rs10000811 | C | T | 0.400 | -0.079 | 0.016 | 7.644E-07 | 24.4 | 0.000 |
| | 110624 | rs75309214 | G | A | 0.099 | 0.202 | 0.043 | 2.219E-06 | 22.4 | 0.000 |
| | 110624 | rs9462875 | G | A | 0.235 | 0.096 | 0.019 | 6.5779E-07 | 24.7 | 0.000 |
| | 110624 | rs2718228 | A | C | 0.540 | 0.105 | 0.021 | 5.173E-07 | 25.2 | 0.000 |
| | 110624 | rs149487444 | C | T | 0.130 | 0.140 | 0.031 | 4.644E-06 | 21.0 | 0.000 |
| | 110624 | rs75933622 | A | G | 0.089 | -0.269 | 0.057 | 2.565E-06 | 22.1 | 0.000 |
| | 110624 | rs147942333 | A | T | 0.075 | 0.415 | 0.080 | 2.168E-07 | 26.9 | 0.000 |
| | 110624 | rs78101397 | C | A | 0.025 | 0.246 | 0.051 | 1.548E-06 | 23.1 | 0.000 |
| | 110624 | rs436772 | T | C | 0.854 | 0.151 | 0.032 | 2.247E-06 | 22.4 | 0.000 |
| | 110624 | rs144495330 | G | A | 0.081 | 0.297 | 0.063 | 2.682E-06 | 22.0 | 0.000 |
| Harmonised SNPs | | | | | | | | | 23.9 | 0.0012 |
| | When assuming a prevalence of 10%, total $R^2$=0.0051 (including 1 proxy SNP) | | | | | | | | | |
| C2: Covid vs population | 2,321,963 | rs148063273 | T | C | 0.008178 | 0.13 | 0.023 | 8.47E-09 | 33.2 | 1.43E-05 |
| | 2,544,875 | rs2109069 | A | G | 0.3084 | 0.033 | 0.005 | 1.68E-12 | 49.8 | 1.96E-05 |
| | 2,572,016 | rs34288077 | G | A | 0.07597 | 0.077 | 0.008 | 1.17E-23 | 100.5 | 3.91E-05 |
| | 2,579,055 | rs4342086 | A | G | 0.3399 | -0.04 | 0.004 | 5.59E-20 | 83.8 | 3.25E-05 |
| | 2,513,309 | rs4801778 | T | G | 0.174 | -0.035 | 0.005 | 1.52E-10 | 41 | 1.63E-05 |
| | 2,586,332 | rs505922 | T | C | 0.6505 | -0.074 | 0.004 | 8.64E-68 | 302.7 | 1.17E-04 |
| | 2,234,283 | rs73062389 | A | G | 0.04899 | 0.162 | 0.009 | 1.54E-65 | 292.3 | 1.3 E-04 |
| Harmonised SNPs | | | | | | | | | 134.2 | 0.0002 |
| | When assuming a prevalence of 5%, total $R^2$=0.00069 | | | | | | | | | |
| D1: Predicted Covid vs predicted or self-reported negative | 38932 | rs10926010 | G | A | 0.189 | -0.170 | 0.036 | 2.762E-06 | 22.0 | 0.001 |
| | 38932 | rs144997841 | A | G | 0.026 | 0.413 | 0.087 | 2.253E-06 | 22.4 | 0.001 |
| | 38932 | rs146540521 | T | C | 0.012 | 0.658 | 0.136 | 1.212E-06 | 23.6 | 0.001 |
| | 38932 | rs115480835 | A | G | 0.029 | 0.513 | 0.110 | 3.141E-06 | 21.7 | 0.001 |
| | 38932 | rs13161855 | A | G | 0.135 | -0.189 | 0.041 | 3.145E-06 | 21.7 | 0.001 |
| | 38932 | rs11952066 | T | C | 0.788 | -0.224 | 0.049 | 4.447E-06 | 21.1 | 0.001 |
| | 38932 | rs72799046 | A | G | 0.058 | 0.319 | 0.064 | 6.3971E-07 | 24.8 | 0.001 |
| | 38932 | rs144932426 | A | G | 0.013 | 0.691 | 0.145 | 1.871E-06 | 22.7 | 0.001 |

| | | | | | | | | | | |
|---|---|---|---|---|---|---|---|---|---|---|
| | 38932 | rs12972001 | G | A | 0.141 | -0.194 | 0.042 | 3.703E-06 | 21.4 | 0.001 |
| Harmonised SNPs | | | | | | | | | 21.7 | 0.0006 |
| When assuming a prevalence of 10%, total $R^2$=0.005 (including 2 proxy SNPs) | | | | | | | | | | |

EAF: effect allele frequency; SE: standard error

SNPs included after data harmonisation with BMI summary data are highlighted in grey

Note: p-value threshold for instruments was reduced to $p < 5 \times 10^{-6}$ for groups A1, C1 and D1

**Supplementary Figure 6 Forest plots of single SNP analysis of the association between COVID-19 and body mass index (BMI): for comparisons A1, A2 and B2 respectively (where there was some evidence of SNP heterogeneity)**

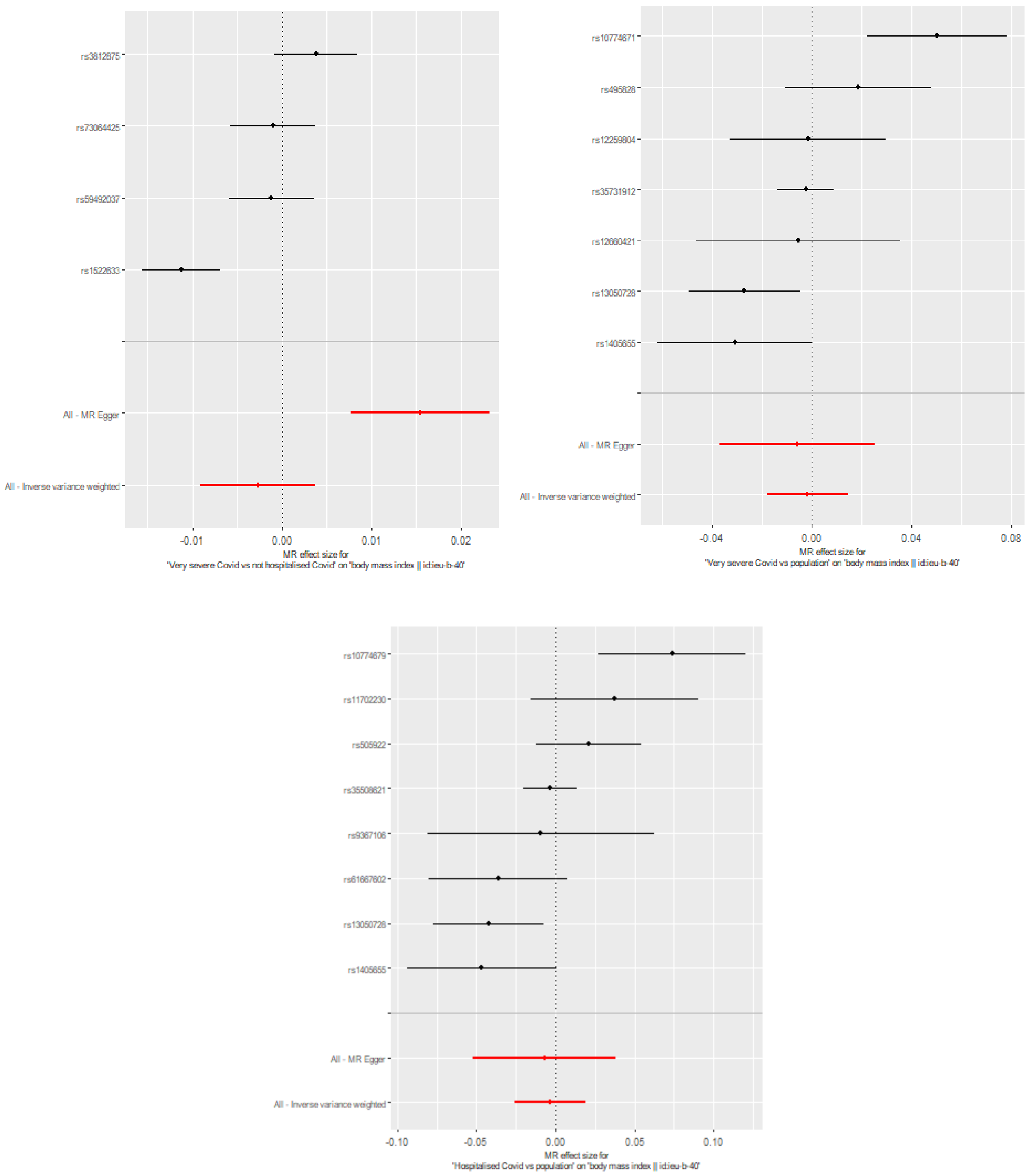


A1: very severe respiratory confirmed COVID-19 vs not hospitalised COVID-19; A2: very severe respiratory confirmed COVID-19 vs population; B2: hospitalised COVID-19 vs population
